# Neuro-Ocular Vasculopathy Associated with Fanconi Anemia: A Clinical-Radiologic Phenotypic Case Series

**DOI:** 10.64898/2026.09.02.26362096

**Authors:** Greer Waldrop, Darshan Pandya, Bryan R. Smith, Ariane Soldatos, Ahmed Abdelhak, Nailyn Rasool, Yair Mina, Christopher Friend, Raphaela Goldbach-Mansky, Kevin Y. Zhang, Calixto-Hope G. Lucas, Kelsey C. Zorn, Sukhman Sidhu, Ari J. Green, Iris Tilton, Samuel J. Pleasure, Neelam Giri, Camilo Toro, Avindra Nath, Michael R. Wilson, Prashanth S. Ramachandran

## Abstract

**Objective:** To characterize the clinical, radiological, and pathological features of Neuro-Ocular Vasculopathy Associated with Fanconi Anemia (NOVA-FA).

**Methods:** Six patients with FA and neurological symptoms were phenotyped using retrospective chart review of clinical notes, neuroimaging, fluorescein angiogram, CSF analysis, and advanced research techniques including extensive infectious testing. Biopsies were reviewed in two patients.

**Results:** NOVA-FA is defined by neurological symptoms, retinal vasculopathy, accumulation of punctate cerebellar lesions and/or large mass-like lesions with surrounding vasogenic edema. Histopathology showed vasculopathy without inflammation. Treatments with immunosuppressants appeared largely ineffective. Imaging and pathology suggest a non-inflammatory small vessel vasculopathy as the primary pathology.

**Interpretation:** NOVA-FA is a recently identified condition that affects a subset of patients with FA and results in significant disability, including death. The etiology remains unknown, although the condition mimics features of the syndrome “retinal vasculopathy and cerebral leukodystrophy” which is due to dysfunction in DNA repair mechanisms. There is currently no known effective treatment.

**Key messages:** *What is already known on this topic:* Fanconi anemia is an inherited DNA-repair disorder, and only scattered case reports and small series have described an acquired neurological and ocular condition in a subset of these patients, and often presumed to be inflammatory. A consistent clinical-radiological definition, an understanding of the underlying mechanism, and evidence to guide treatment have been lacking.

*What this study adds:* This series of six patients provides the most detailed clinical, radiological and pathological characterization of the syndrome to date, proposes the name Neuro-Ocular Vasculopathy Associated with Fanconi Anemia (NOVA-FA) with provisional diagnostic criteria, and shows that histopathology, CSF and serum biomarkers, infectious testing and treatment response together point to a non-inflammatory small vessel vasculopathy rather than a primary immune-mediated or infectious process, closely mirroring the DNA-repair disorder RVCL.

*How this study might affect research, practice or policy:* The findings argue against reflexive immunosuppression, support earlier recognition of NOVA-FA through defined criteria, and reframe it as a DNA-repair-driven vasculopathy, motivating natural-history and mechanistic studies and the evaluation of vascular-targeted therapies as FA life expectancy rises and this phenotype becomes more common.

**Social Media Summary If Published:** None of the co-authors have a X handle that we would like to be tagged. There are scant case reports about this emerging neurologic syndrome in patients with Fanconi Anemia. This study contains the most robust neurological phenotyping of these patients and explores in-depth evaluations for molecular presence of inflammation.

Draft: A novel progressive neuro-ocular vasculopathy in Fanconi Anemia (NOVA-FA) presents with enhancing brain lesions and retinal changes. Likely due to microvascular DNA repair failure, not inflammation. #FanconiAnemia #Neurovascular #RareDisease

## Introduction

Fanconi anemia (FA) is a rare and diverse inherited, DNA-repair disorder associated with mutations in up to 22 different genes. These genes contribute to the FA pathway, a series of proteins essential for DNA repair and maintenance. Consequently, the hallmark of FA is a compromised ability to repair DNA interstrand crosslinks (ICs). This impairment leads to an accumulation of ICs in the genome, resulting in disrupted cell division and DNA replication, heightened genetic instability, bone marrow failure, and an elevated risk of malignancies.^1^ Approximately 75% of FA patients exhibit congenital birth defects and structural abnormalities, including central nervous system (CNS) and skull base developmental anomalies in a specific subgroup.^2–5^ One of the most prevalent manifestations is acquired pancytopenia, often necessitating bone marrow transplantation.

FA shares several similarities to other DNA repair disorders including, developmental abnormalities, bone marrow failure and predisposition to cancer. In the past 20 years, there have been case reports and small cases series that have described a subset of FA patients exhibiting a shared phenotype of an acquired neurological and ocular condition^1–11^.^6–16^ Although this syndrome has been described previously with the general term, “Fanconi Anemia Neurologic Syndrome”, we propose the term Neuro-ocular Vasculopathy Associated with FA (NOVA-FA) to described with more precision this emerging phenotype, characterized by progressive small vessel vasculopathy. NOVA-FA reflects a manifestation of FA-related DNA repair dysfunction and highlights the convergence of neurovascular and ocular pathology in a subset of FA patients. Here, we describe the clinico-radiological phenotype of this condition, a proposed and provisional diagnostic criterion, and provide insights into the underlying etiology and pathophysiology of NOVA-FA (Figure 1).

**Figure 1:**
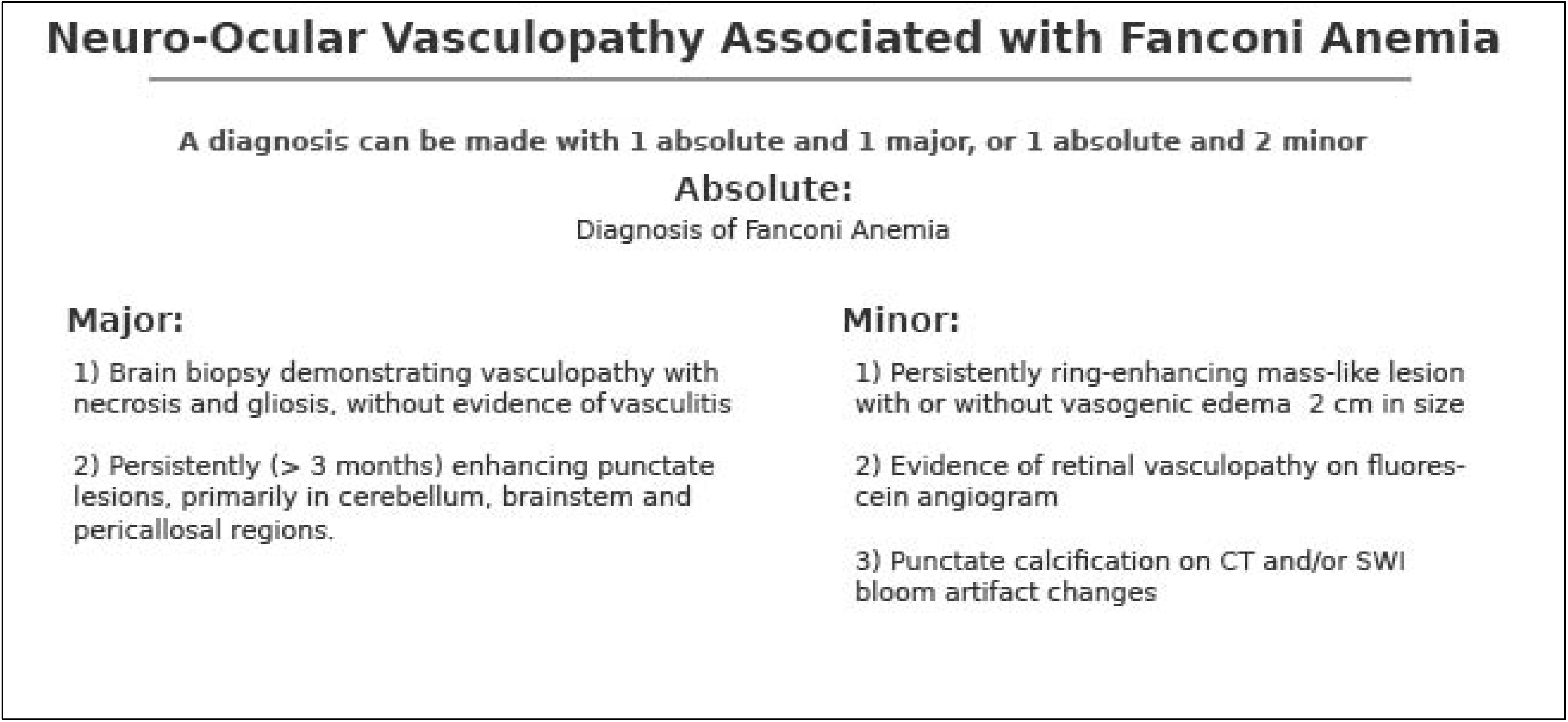
Proposed Provisional Diagnostic Criteria for Neuro-Ocular Vasculopathy Associated with Fanconi Anemia.

## Methods

### Patient Cohort

Patients were enrolled in the retrospective cohort study if they had a diagnosis of FA and neurological symptoms and abnormal MRI brain results not otherwise explained by another neurological condition. Patients were frequently referred from the FA Research Fund, a non-profit organization. Participants were consecutively enrolled from clinical practice within the UCSF Multiple Sclerosis (MS) and Neuroinflammation Center or from referrals to the National Institutes of Health (NIH) Clinical Center via the UCSF/NIH Investigation and Treatment of Undiagnosed Neuroinflammatory Diseases study (U01NS120836). All research was approved at local institutional review boards (UCSF IRB 13-12236) and included appropriate clauses for data sharing.

Enrolment included complete medical records review (retrospective and prospective) including demographic information, clinical history, genetic results, serum and cerebrospinal fluid (CSF) laboratory results, ophthalmology reports, radiology reports and images. Demographics, important clinical features, including neurological phenotype, laboratory testing, and radiological features were abstracted into an online natural history registry.

All MRI scans were reviewed by a board-certified neurologist (GW) for further qualitative characterization and quantification of lesions, by neuroanatomical location. The following classification, modified from other vasculopathy descriptions,^17^ was used to describe types of T_2_-hyperintense white matter lesions identified with neuroimaging: (i) punctate lesions without enhancement; (ii) punctate lesions with enhancement; (iii) large mass-like lesions with surrounding edema; (iv) large mass-like lesions without surrounding edema.

For patients who underwent brain biopsy, formalin fixed paraffin embedded brain biopsy tissue underwent re-review with a neuropathologist at the NIH and The Johns Hopkins University.

Additionally, consent was obtained for research-based testing using surplus biological specimens (i.e. serum, CSF, tissue) from already collected clinical samples. The tests included metagenomic next generation sequencing (mNGS), CSF neopterin quantification, rodent brain immunostaining, Luminex-based cytokine analysis, transcriptional analysis of 28 type I interferon-response genes using Nanostring technology^18^, CSF protein abundance levels were measured using Olink HT Explore, proximity extension assay platform^19^ and intensity□based retinal arteriolar visualization in optical coherence tomography (I□bRAVO)^20^. Methodology for these assays have been described previously and can be found in the Supplementary Appendix.

## Results

Six participants were enrolled. Two additional patients were identified and met criteria for diagnosis but declined consent for research. For three consented patients, additional research-based testing from clinically collected biospecimens was performed. Four patients have been described in a prior publication^8^, but additional in-depth neurological phenotyping, imaging review, research-based testing, and longitudinal data are included here. On average there was 5 years of follow up time (range 3-9 years) after diagnosis of NOVA-FA, half of the patients were male sex (male gender), and all but one patient were of white race (83%). Four unique FA genes were identified (FANCD-2, FANC-J, FANC-F, FANC-C), and one patient was diagnosed based on a chromosomal breakage test and did not have an abnormal FA gene identified despite whole exome testing. Most patients were diagnosed with FA within the first year of life (n=4, 67%). None of the patients had neurological symptoms at the time of FA diagnosis. Three patients (50%) required bone marrow transplant. Summary results are presented in Table 1, and individual patient-level data are presented in Supplemental Table 1.

**Table 1:** Demographic, Clinical, Laboratory and Radiological Characteristics of Patients Neuro-Ocular Vasculopathy Associated with Fanconi Anemia (NOVA-FA) (n=6)

|  | **N(%)** |
| --- | --- |
| Male Sex | 3 (50) |
| Race/Ethnicity |  |
| American Indian/Native Alaskan | 0 (0) |
| Asian | 1 (17) |
| Black or African American | 0 (0) |
| Native Hawaiian or Other Pacific Islander | 0 (0) |
| White, Non-Hispanic | 5 (83) |
| More than one race | 0 (0) |
| Unknown, not reported | 0 (0) |
| Years of Follow-up Since FANS Diagnosis (Mean (SD)) | 5 (2) |
| **Fanconi Anemia (FA) History** |  |
| Number of Unique FA Genes Identified | 4 |
| Age in Years at FA Diagnosis (Mean (SD)) | 6 (8) |
| FA Diagnosis within first year of Life | 4 (67) |
| Neurological Symptoms at FA Diagnosis | 0 (0) |
| Bone Marrow Transplant | 3 (50) |
| Fanconi Anemia Neurological Syndrome |  |
| Clinical Characteristics at FAANS Diagnosis |  |
| Age at FAANS Onset (Mean (SD)) | 21 (7) |
| Years from FA Diagnosis to FAANS Onset (Mean (SD)) | 15 (6) |
| History of Neurological Symptoms | 5 (83) |
| Initial Neurological Symptoms |  |
| Ataxia and Hand Dysmetria, Subacute onset | 1 (17) |
| Hemiplegia, Acute Onset | 2 (33) |
| Hemiplegia, Subacute Onset | 1 (17) |
| Vertigo, Acute Onset Persistent | 1 (17) |
| Vertigo, Acute Onset Transient | 1 (17) |
| Seizures | 3 (50) |
| Retinopathy* | 5 (100) |
| **FAANS Clinical Course** |  |
| Asymptomatic/Mild intermittent symptoms | 1 (17) |
| Focal neurological attacks only | 1 (17) |
| Focal neurological attacks with interval progression | 1 (17) |
| Progressive symptoms without focal neurological attacks | 3 (34) |
| mRS at last clinical encounter |  |
| 1 | 1 (17) |
| 2 | 1 (17) |
| 3 | 1 (17) |
| 4 | 0 (0) |
| 5 | 1 (17) |
| 6 | 2 (33) |
| **CSF Results** |  |
| Pleocytosis | 0 (0) |
| Elevated Protein (per clinical testing range) | 5 (83) |
| Elevated Protein > 100 mg/dL | 5 (83) |
| Elevated IgG Index | 0 (0) |
| JCV PCR Positive* | 0 (0) |
| **Radiological Characteristics** |  |
| At least one Mass-like Lesion with gadolinium enhancement | 4 (67) |
| Associated with Vasogenic Edema*** | 3 (75) |
| Punctate T2 lesions with gadolinium enhancement | 6 (100) |
| Cortex and Subcortical Regions | 5 (83) |
| Brainstem | 6 (100) |
| Cerebellum | 5 (83) |
| Calcifications on CT Head | 5 (100)**** |
| Evidence of SWI Microhemorrhages | 4 (67) |
| Spinal Cord Involvement | 4 (67) |
| Cervical** | 1 (25) |
| Thoracic** | 3 (75) |
| **Treatment Trials** |  |
| Anakinra | 2 (33) |
| Cyclosporin | 1 (17) |
| Intravenous Immunoglobulin | 2 (33) |
| Methylprednisolone | 3 (50) |
| Oral Glucocorticoids | 3 (50) |
| Rituximab | 1 (17) |
| Sirolimus | 2 (33) |
| Tocilizumab | 1 (17) |

\*4 patient tested for CSF JCV PCR, 5 patients tested with formal florecin angiograms
\*\*Among those with spinal cord involvement
\*\*\*among those with mass like lesions
\*\*\*\* 5 patients had CT Head evaluations for review
Abbreviations: SD standard deviation , mRS modified rankin score, CSF Cerebrospinal Fluid mg milligram, dl deciliter, IgG Immunoglobulin G, JCV John Cunningham Virus, PCR Polymerase C T2 transverse relaxation time, mm milimeter, SWI Suseptibility Weighted Imaging

### NOVA-FA Presentation

The average age at NOVA-FA symptom onset was 21 years old (range: 13-32 years), with an average of 15 years (range: 7-23 years) between the diagnosis of FA and the onset of NOVA-FA. The majority of patients, in retrospect, had a history of neurological symptoms prior to the onset of NOVA-FA (n=5, 83%) including complex migraine, neurocognitive deficits, clumsiness, or chronic vertigo. All patients had evidence of retinopathy at the time of the NOVA-FA diagnosis. (Table 1, Supplemental Table 1).

### Neurological Syndrome

Both acute attacks, subacute progressive, and mild-intermittent presentations occur within NOVA-FA, and these categorizations are not mutually exclusive. Half of the patients presented acutely with their first symptoms of NOVA-FA. Overall, four of the six patients experienced an acute attack during the disease course, as one patient’s acute attack was preceded by a subacute presentation. Seizures occurred in fifty percent of patients, including patients with acute and subacute presentations. One of the six patients experienced a non-progressive clinical course with mild, episodic vertigo.

The acute attacks include vertigo and hemiplegia, which were preceded by a fall or physical trauma in two cases. Recurrent, acute attacks occurred but not in every patient. Clinical stability between attacks, and clinical progression between attacks were both reported. One patient died due to cerebral edema and herniation during an acute attack. All the acute attacks were associated with mass-like lesions, with or without vasogenic edema, as described in the Imaging Results section. For the patients with who did not experience any acute attacks, the presentations included subacute ataxia and subacute hemiplegia.

### Visual Syndrome

Five of the six patients demonstrated significant deficits in visual acuity. Visual acuities in a single eye ranged from 20/20 to finger counting. Formal visual acuities were 20/20, 20/40, 20/160, 20/200 bilaterally, for four patients respectively and for two patients there was asymmetry between the eyes with 20/50 in one eye and finger counting only in the other.

### Radiological Course

NOVA-FA patients have characteristic changes on MRIs of the brain and, to a lesser degree, the spinal cord. A qualitative summary of radiological findings is reported in Supplemental Table 3. All NOVA-FA patients developed punctate T_2_-hyperintense brain lesions with persistent contrast enhancement, most commonly (but not exclusively) in the cerebellum. These punctate lesions were associated with T1 central hypointensity. Some lesions exhibited contrast enhancement lasting several years. The number of punctate lesions varied significantly between patients. For example, at the end of follow-up for all patient the range of enhancing punctate lesions in the cerebellum ranged from 2 – 60, brainstem 1-5, and bilateral supratentorial hemispheres 2 – 80, estimated by visual inspection.

In addition to these features, imaging features included:

1. Calcifications
2. Susceptibility weighted imaging (SWI) changes
3. Large mass-like lesions with and without surrounding edema
4. Diffusion restriction
5. Atrophy, both focal or generalized
6. Spinal cord lesions

Four of the six patients developed multiple, large mass-like lesions. (Supplemental Figure 2). Three of these patients developed two lesions and an additional patient developed a third, resulting in 9 total mass-like lesions. These mass-like lesions all demonstrated complete ring, gadolinium-contrast enhancement. Three lesions developed in the corpus callosum, and the lesions were located in the right frontal lobe (n=2), left frontal lobe (n=1), left centrum semiovale (n=1), left parietal lobe (n=1), and left occipital lobe (n=1). Serial imaging demonstrated the slow growth over several months of one of these lesions. Three of these patients developed vasogenic edema associated with at least one of their mass-like lesions. Large lesions with severe vasogenic edema led to midline shift, falcine and uncal herniation.

High dose glucocorticoids were administered to all patients with mass-like lesions and was associated with temporary improvement in vasogenic edema when administered with a return of edema on early steroid wean or cessation. Glucocorticoid treatment failed to reduce the size or contrast enhancement of these mass-like lesions. Independent of treatment, mass-like lesions tended to involute over 2-3 months with subsequent development of focal atrophy and gliosis and slow resolution of contrast enhancement. Vasogenic edema slowly resolved during this time frame as well. The mass-like lesions demonstrated persistent diffusion restriction over several months.

Figure 2 demonstrates the course of a single patient over four years with mass-like lesions with and without vasogenic edema and the accumulation of T_2_-hyperintense, gadolinium enhancing lesions and SWI signal voids. Figure 3 demonstrates the progression of one patient’s imaging findings over 7 years, notably the accumulation of punctate, contrast enhancing lesions in the cerebellum, and the accumulation of the SWI changes sparing the cerebellum. Indeed, SWI signal voids occurred in 5 of the six patients, largely sparing the cerebellum, despite the predilection of the cerebellum for the punctate T_2_-hyperintense lesions.

**Figure 2:**
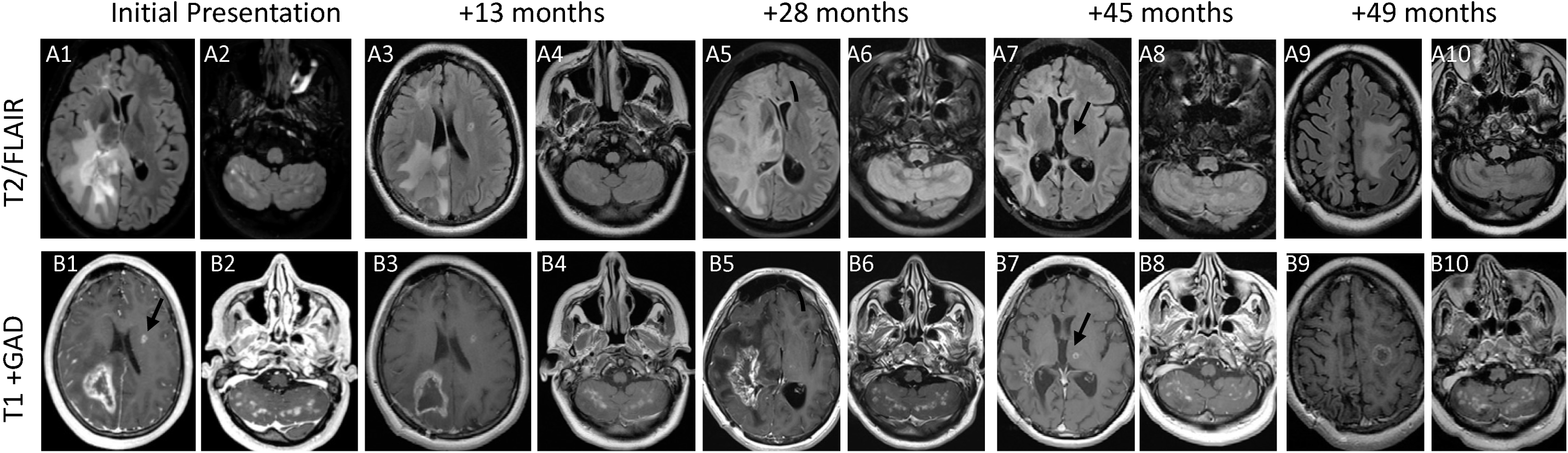
Representative images of MRI Brain with recurrent gadolinium enhancing mass-like lesions with and without surrounding edema and accumulating punctate gadolinium enhancing lesions in a single patient living with NOVA over 49 months. This patient noted subacute dragging of left leg, and clumsiness of the left hand for three months prior to afterwards episode of transient (lasting 30 minutes) hyperacute left hemiplegia, immediately after a fall while playing basketball. MRI revealed a rim-enhancing lesion with mass-effect and surrounding oedema in the right parietal white matter and smaller enhancing lesion without surrounded edema in the left frontal lobe (black arrow) (**A1**: Fluid attenuated inversion recovery (FLAIR) sequence; **B1**: gadolinium-enhanced T_1_-weighted sequence). Numerous FLAIR and enhancing lesions were noted bilateral cerebellum ( **A2**:FLAIR sequence; **B2**: gadolinium-enhanced T_1_-weighted sequence) Thirteen months later, after trial of glucocorticoids and a generalized seizure there was mildly increased size of the right parietal lesion with decreased size and mass effect of surrounding edema with persistent enhancement of prior supratentorial (**A3**: FLAIR sequence; **B3**: gadolinium-enhanced T_1_-weighted sequence) and infratentorial lesions (**A4**:FLAIR sequence; **B4**: gadolinium-enhanced T_1_-weighted sequence). Twenty eight months after initial presentation patient developed nausea, vomiting, vertigo. MRI revealed a left periventricular rim enhancing lesion with surrounding edema and mass effect (**A5**: FLAIR sequence; **B5**: gadolinium-enhanced T_1_-weighted sequence) and increase in quantity of infratentorial enhancing lesions (**A6**: FLAIR sequence; **B6**: gadolinium-enhanced T_1_-weighted sequence). Forty-five months from initial presentation, on treatment with Sirolimus, there was improvement of size, edema and degree of enhancement of left periventricular lesion but new lesion in the left thalamus without surrounding edema (black arrow) (**A7**: FLAIR sequence; **B7**: gadolinium-enhanced T_1_-weighted sequence) and stable quantity of enhancing infratentorial lesions (**A8**: FLAIR sequence; **B8:** gadolinium-enhanced T_1_-weighted sequence). Forty nine months from initial presentation patient developed acute left hemiparesis and MRI revealed a rim-enhancing lesion with surrounding edema in the right frontal lobe **A9**: FLAIR sequence; **B9**: gadolinium-enhanced T_1_-weighted sequence and stable to mildly increased quantity of enhancing infratentorial lesions (**A10**: FLAIR sequence; **B10**: gadolinium-enhanced T_1_-weighted sequence). Abbreviations: T2/FLAIR, T_2_-weighted sequence/Fluid-Attenuated Inversion Recovery , T1+ GAD T1 weighted Sequences with Gadolinium Enhancement

**Figure 3:**
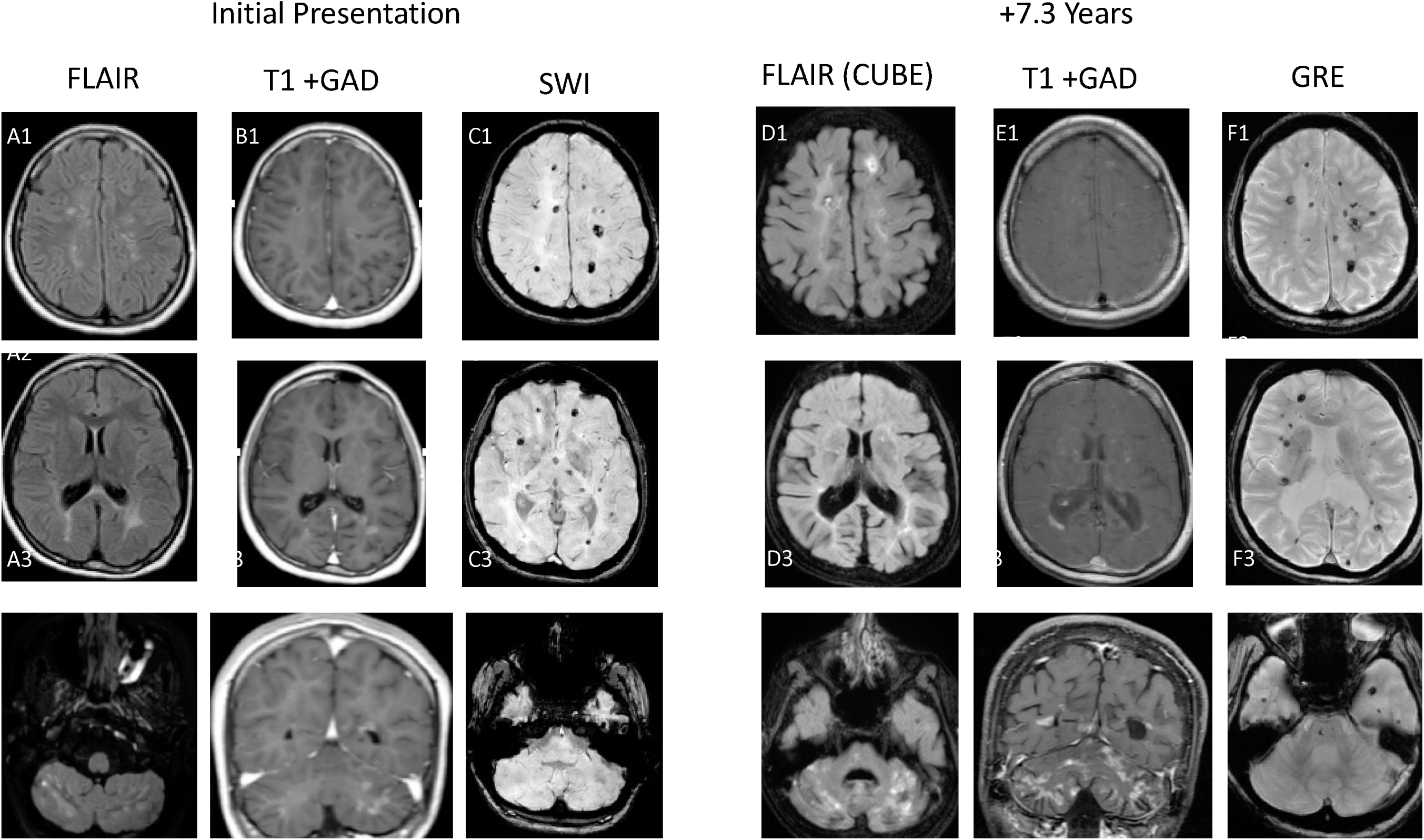
Representative images of MRI Brain demonstrating primarily infratentorial accumulation of punctate gadolinium enhancing lesions and accumulation of primarily supratentorial hemosiderin deposits in a single patient living with NOVA over seven years. This patient presented with subacute right hand weakness and difficulty with balance and over seven years continued to experience ataxia, cognitive impairment and seizures. MRI Brain at most recent clinical encounter (Panels D-F) compared to initial MRI Brain (Panels A-C) seven years and 4 months prior, demonstrates accumulation of numerous punctate FLAIR (A, D Panels) and gadolinium enhancing (B, E Panels) lesions in bilateral cortical white matter (1), bilateral posterior periventricular subcortical white matter (2) and most prominently in bilateral brainstem and cerebellum (3). Additionally, accumulation of numerous hypointense SWI signal (C, F Panels) in bilateral cortical white matter (1), bilateral posterior periventricular subcortical white matter (2) and brainstem, sparing the cerebellum (3).Abbreviations: FLAIR, Fluid-Attenuated Inversion Recovery, T1+ GAD T_1_ weighted Sequences with Gadolinium Enhancement, SWI, Susceptibility-weighted imaging FLAIR CUBE, Fluid-Attenuated Inversion Recovery with 3D fast spin echo sequence, GRE, Gradient Echo Sequences

Of the five patients with CT scan images of the head, all demonstrated evidence of calcifications. Though there was some correlation with SWI bloom artifact with calcific lesions on CT scan, not all SWI lesions correlated with areas of calcification, and we therefore could not comment on whether all SWI changes represent micro-calcification or hemosiderin deposition. Mass like lesions developed in patients with both high and low burden of punctate, enhancing T2 lesions.

Short segment, gadolinium enhancing T_2_-hyperintense intramedullary spinal cord lesions without clear track predominance were identified in four of the six patients (Supplemental Figure 2). Five of the six patients demonstrated T1 hypointensities in the corpus callosum. Focal atrophy was evident for all patients at the location of longstanding punctate lesions, spinal cord lesions and mass-like lesions. Qualitatively, it was observed that patients with a high burden of punctate lesions developed more generalized cerebellar and cerebral atrophy (Supplemental Figure 3).

### Brain Pathology

Histopathological examination from brain biopsies in two patients revealed parenchymal necrosis, abundant macrophages and reactive gliosis, with one patient demonstrating calcifications.

Vascular changes were evident in brain tissue from one patient, characterized on Verhoeff-Van Gieson (VVG) staining with a lack of elastic lamina around the hyalinized vessels. Trichrome staining demonstrated thick-walled vessels and showed luminal narrowing and occlusion in multiple small vessels, both within the areas of necrosis and the adjacent brain parenchyma.

The other patient’s brain biopsy demonstrated perivascular lymphocytic CD4+ infiltrates but no evidence of vasculitis and no evidence of luminal narrowing. JC and BK virus staining was negative, as was staining for other microorganisms. Luxol fast blue staining did not demonstrate demyelination. Detailed pathology reports and images can be found in Supplemental Table 3 and Supplemental Figure 5.

### Clinical CSF Studies

All six patients had clinical CSF analysis. None of the patients demonstrated a pleocytosis or elevated IgG index. Five patients (83%) demonstrated elevated CSF protein, all of which were above 100 mg/dL. Four patients were tested for JC virus (JCV) by a CSF JCV PCR. All PCRs were negative. One patient was tested for CSF angiotensin converting enzyme (ACE) which was elevated (4.7 U/L, Normal range: 0.0 - 2.5 U/L). Supplemental Table 1 reports comprehensive CSF profiles.

Two patients underwent CSF cytokine analysis using at ARUP which includes 13 cytokines: soluble Interleukin (IL)-interleukin2 receptor, IL-interleukin12, Interferon-gamma, IL-4, IL-5, IL-10, IL-13, IL-1 beta, IL-6, IL-8, tumor necrosis factor-alpha, IL-2, and IL-17. One patient had normal cytokine levels, and the other patient had elevated IL-6 (17 pg/mL, normal<0.5 pg/mL).

### Retinal Findings

Five patients had a formal neuro-ophthalmological evaluation. All had evidence of a retinal vasculopathy, demonstrating telangiectasia and tortuous vessels. In all five patients, fluorescein angiogram demonstrated abnormal vessels with evidence of leakage and capillary non-perfusion. Ocular coherence tomography (OCT) demonstrated thinning of the retinal nerve fiber layer. None of the patients demonstrated evidence of vasculitis. (Figure 4)

**Figure 4:**
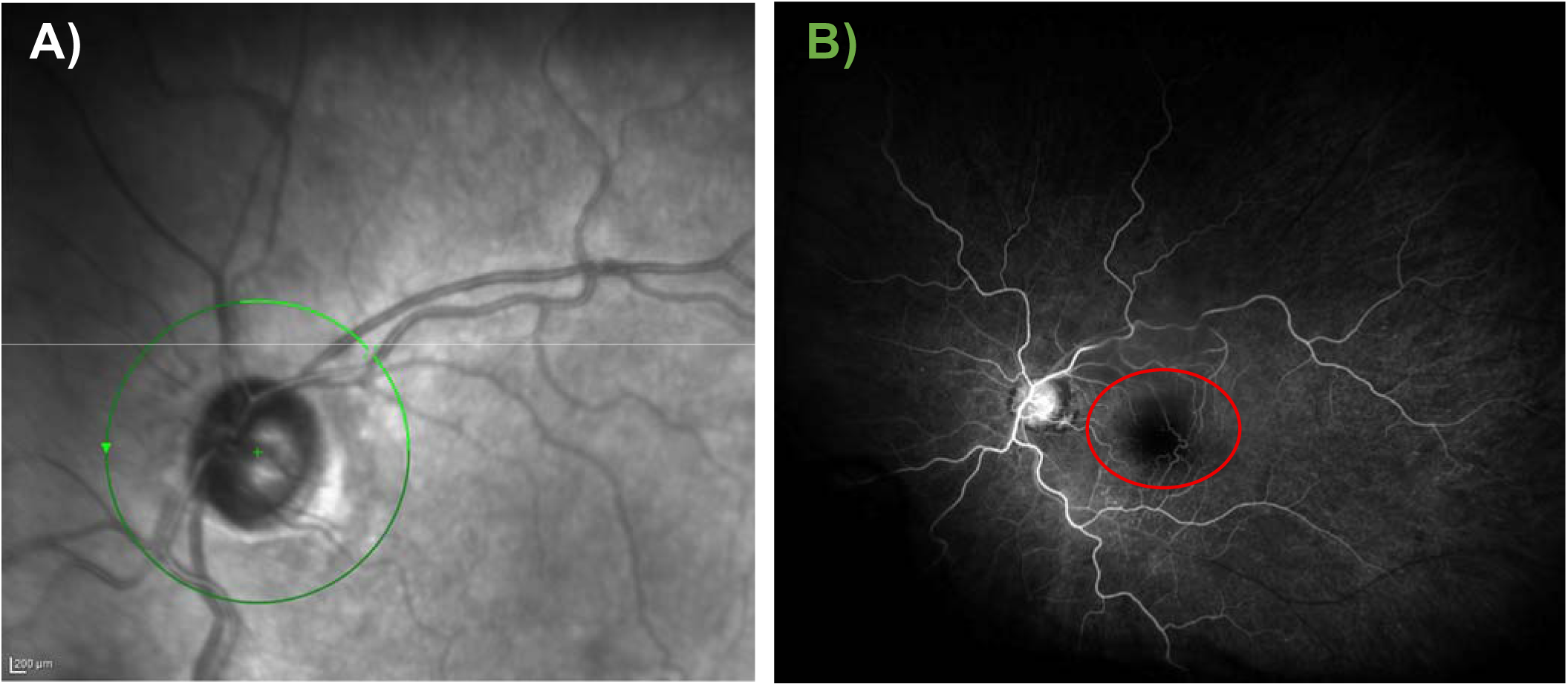
Representative images of florescence angiogram and funduscopic images demonstrating from a single patient living with NOVA. A) Funduscopic image illustrating vascular telangectasias surrounding optic nerve B) Florescence Angiogram illustrating Capillary nonperfusion of macula with enlargement of the foveal avascular zone (red circle)

### (I□bRAVO) Findings

Two patients underwent formal vessel wall measurement with I-bRAVO. Individual lumen diameters for both patients were narrower than the mean lumen diameters from healthy control cohort and from cerebral small vessel disease (CSVD) cohort. Individual inner wall diameters were wider than healthy controls or CSVD in one NOVA-FA patient, but thinner in the other NOVA-FA. Individual outer wall diameters were thinner in both NOVA-FA patients than the healthy control and CSVD cohorts. Complete reports including all measurements available in Table 2.

**Table 2:**
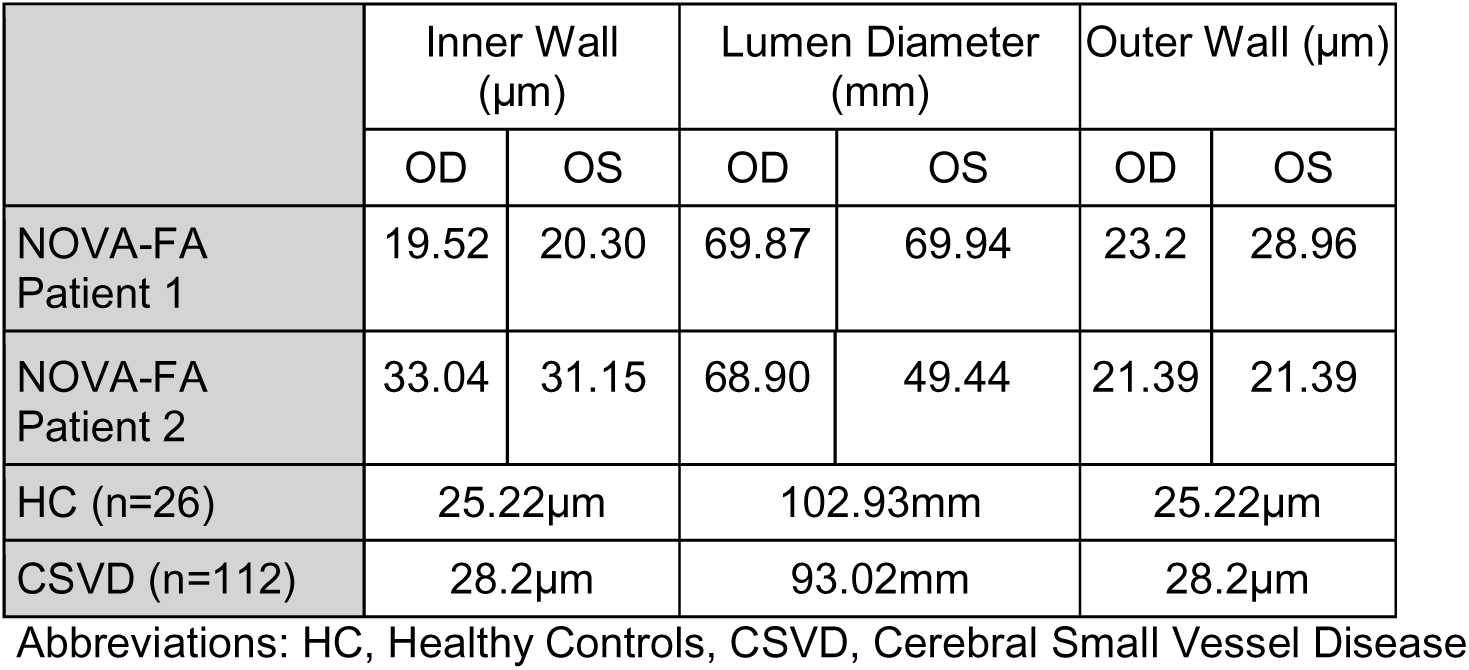
Summary of Vessel Wall Measurements in Two Patients with NOVA-FA, Healthy Controls and Patients with Cerebral Small Vessel Disease.

|  | Inner Wall (μm) |  | Lumen Diameter (mm) |  | Outer Wall (μm) |  |
| --- | --- | --- | --- | --- | --- | --- |
|  | OD | OS | OD | OS | OD | OS |
| NOVA-FA Patient 1 | 19.52 | 20.30 | 69.87 | 69.94 | 23.2 | 28.96 |
| NOVA-FA Patient 2 | 33.04 | 31.15 | 68.90 | 49.44 | 21.39 | 21.39 |
| HC (n=26) | 25.22μm |  | 102.93mm |  | 25.22μm |  |
| CSVD (n=112) | 28.2μm |  | 93.02mm |  | 28.2μm |  |

Abbreviations: HC, Healthy Controls, CSVD, Cerebral Small Vessel Disease

### Treatment and Outcomes

With the exception of the patient with mild, intermittent, non-progressive vertigo, all patients were treated empirically with various combinations of steroid-sparing immunomodulatory therapy. The following treatments were trialed: anakinra, cyclosporin, intravenous immunoglobulin, mycophenolate mofetil, rituximab, sirolimus, and tocilizumab. None of these treatments, or combinations thereof, were associated with amelioration of progressive symptoms or remission of new acute attacks. The only exception to this was trend was that one patient treated concurrently with anakinra and sirolimus had a stable clinical course for more than 3 years, but other patients on anakinra alone (n=1) had breakthrough disease. As mentioned previously, Intravenous methylprednisolone was administered to all patients with acute presentations of mass-like lesion, which resulted in improvement of vasogenic edema without changes to enhancement pattern.

With the exception of one patient, who has been lost to follow-up we have updated functional status within a year of this publication. At last follow up, the majority of patients (n=2) walked independently (modified Rankin scale (mRS) <4). One was unable to walk unassisted (mRS: 4), and one patient was severely disabled requiring constant nursing care (mRS: 5). Two patients died due to complications of a severe acute attack with cerebral herniation from mass-like lesions attributed to NOVA-FA (mRS:6). Of the two patients who died, one died while receiving hospice care following a decision to transition to comfort-focused care due to severe neurologic disability resulting from cerebral herniation of a left frontal lesion. The second patient died in the hospital from pneumonia, which developed secondary to severe neurologic deficits sustained from cerebral herniation of a right colossal lesion which progressed over the course of a year. Representative images from the most recent neuroimaging obtained prior to death for both patients are shown in Figure 5.

**Figure 5:**
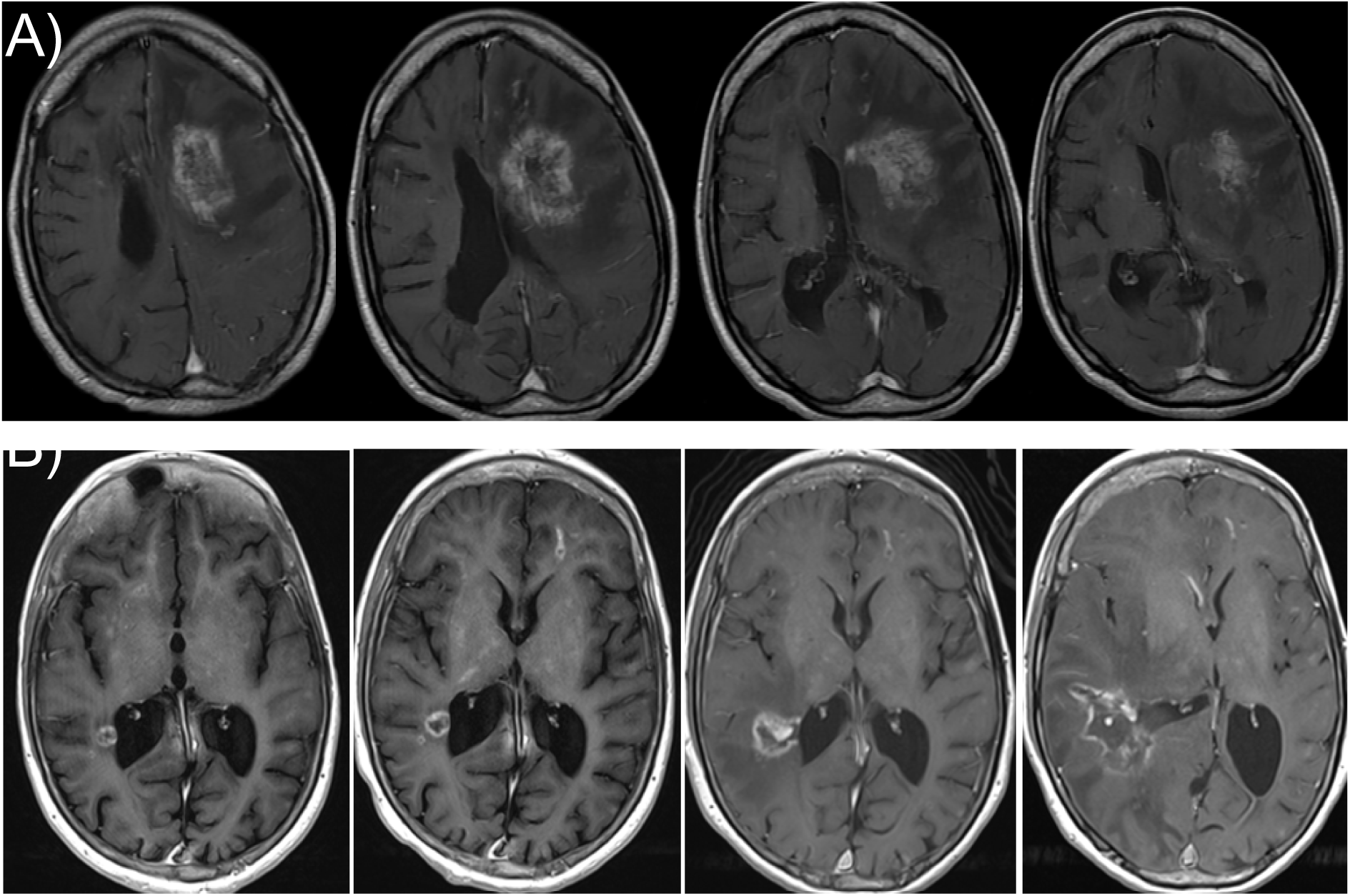
Representative images of cerebral herniation secondary to mass lesions attributed to NOVA-FA. A) A series of T1 weighted sequence with gadolinium contrast images from the same scan illustrating the extent of a left frontal subcortical mass-like lesions with vasogenic edema, mass-effect and falcine herniation occurring prior to death while in hospice care. B) A series of T1 weighted sequence with gadolinium contrast images documenting progression over 11 months of a right posterior collosal mass like lesion with development of vasogenic edema, mass effect and falcine and uncal cerebral herniation. The right-most image occurring prior to in-hospital death.

### Research-Based Testing

For the three patients who completed additional research testing, no viral, bacterial, fungal or parasitic pathogens were identified by CSF mNGS. Two patients were tested for CSF neopterin, and both were not elevated, suggestive of no cellular immune activation. One patient’s CSF was tested for type 1 interferon score measured using Nanostring technologies, and did not demonstrate a high interferon signaling. CSF rodent brain immunostaining with CSF IgG at 1:4 dilution failed to identify a consistent pattern of staining suggestive of an anti-neural antibody across three patients’ CSF samples. (Supplemental Table 4).

Despite small sample sizes, protein abundance assay comparisons between NOVA-FA patients (n=3) and biopsy-confirmed primary CNS vasculitis (n=3), identified 118 statistically significantly differentially abundant CSF proteins. The proteins with the largest differences, by absolute log fold change, included interlukin-6 (decreased abundance in NOVA-FA) and DNA cell cycle proteins increased in NOVA-F (Centromere Protein F). Over-representation analysis demonstrated differences in vascular endothelial growth factor signaling and complement activation (Supplemental Figures 6 and 7).

## Discussion

NOVA-FA is an acquired syndrome that develops in a subset of FA patients typically in their teens to early 20s. These patients may or may not have a history of bone marrow transplantation. There was a significant spectrum of underlying disease activity ranging from mild symptoms with minimal lesion burden and slow rate of lesion accumulation, to severe disease from high lesion burden, high rate of accumulation and subsequent cerebral and cerebellar atrophy. All patients demonstrate evidence retinopathy. Fundamentally though, FA-NOVA presents, neurologically, as a progressive neurological condition with the possibility of superimposed acute attacks. NOVA-FA can lead to weakness, spasticity, epilepsy, vision loss, ataxia, and even death. Acute attacks can manifest with large mass-like lesions with severe vasogenic edema, ultimately leading to brain herniation and death in some cases. Based on these unifying features, we proposed a provisional diagnostic criteria for this syndrome (Figure 1) with the goal of assisting treating physicians as they attempt to differentiate this rare condition from the wide variety of other etiologies that can present with neurologic symptoms and contrast-enhancing brain lesions.

Despite extensive clinical and research-based testing there was scant evidence of a primary immune-mediated process underlying NOVA-FA. Consistent with this, several patients were treated with significant immunosuppression that failed to clearly halt disease progression. While high dose steroids may have helped transiently improve the degree of vasogenic oedema surrounding large mass-like lesions, steroids were not clearly associated with resolution of contrast-enhancement or reduction in the size of the primary lesion. Mild CSF IL-6 elevation on the cytokine analysis of one patient, and microglial/macrophage activation seen on brain biopsy, may represent innate immune activation, but whether this is a primary driver of injury or a non-specific response to an alternate type of pathology is far from clear. In addition, extensive clinical and research testing failed to identify an infectious cause.

Instead, the clinical data, brain pathology and additional research testing of the CSF and retinal vessels supported that the likely mechanism of injury for NOVA-FA is a small vessel vasculopathy. We postulate that this obliterative vasculopathy leads to ischemia, necrosis, gliosis and focal axonal loss. Persistent contrast enhancement and elevated CSF protein may simply be markers of a permeable blood brain barrier in this setting. It is unclear what the mechanism is for the development of mass-like lesions, but this may represent neovascularization, similar to large lesions seen in radiation necrosis, another DNA damage disorder.^21^

Indeed, NOVA-FA shares a striking phenotypic similarity with Retinal Vasculopathy with Cerebral Leukodystrophy (RVCL), another DNA damage repair disorder.^22,23^ RVCL is a monogenic small vessel disease caused by mutations in the *TREX1* gene, which encodes a DNA exonuclease involved in DNA repair. Pathogenic *TREX1* variants result in mis-localization and functional impairment of the protein, leading to accumulation of unrepaired double-stranded DNA breaks, genomic instability, and cellular senescence. Importantly, RVCL is not considered to be an interferonopathy.^24^

The underlying genetic defect in FA impairs the repair of DNA interstrand crosslinks, promoting chronic DNA damage. We postulate that endothelial cells at the blood-brain barrier are particularly susceptible to reactive oxidative damage. These cells typically undergo senescence rather than apoptosis and can exhibit progressive dysfunction, vascular inflammation, and remodelling, particularly within the retina and CNS^12,13^. Both disorders (NOVA-FA and RVCL) demonstrate a progressive small vessel vasculopathy with blood-brain barrier permeability and superimposed neurodegeneration, without evidence of a dominant systemic inflammatory signature. Supplemental Figure 8 compares genetic, clinical and medication trials between RVCL and NOVA-FA. With increases in FA life expectancy and the onset of NOVA-FA in adolescence and early adulthood, we may be witnessing the emergence of a novel phenotypic subtype of FA. It remains unclear whether certain exposures, such as bone marrow transplantation or radiation, may trigger or accelerate this process.

Vascular endothelial growth factor (VEGF) inhibitors or Boswellia may be a treatment option for NOVA-FA patients with life threatening, mass-like lesions with vasogenic edema and midline shift, but this suggestion is purely based on comparisons to use of this treatment strategy in radiation necrosis.^25,26^ At present, we would recommend high dose steroids for patients with severe vasogenic edema from mass-like lesions in life threatening situations, with appreciation that this will likely not alter the underlying disease course.

## Conclusion

NOVA-FA is an entity characterized by cerebral and retinal vasculopathy affecting patients with FA and phenotypically carries a striking resemblance to RVCL, another genetic condition related to DNA damage and repair. Despite characteristic MRI features that include persistent contrast enhancement, there is little pathological, biomarker and therapeutic response evidence for an inflammatory process. More research is needed to further understand the natural history of this syndrome and its pathophysiology.

## Acknowledgements

The authors would like to thank the patients and their families for participation in this nascent and critical stage for the understanding of this rare disease. We would also like to thank the Fanconi Cancer Foundation, formerly FA Research Foundation for their dedication to these patients and efforts towards advancing research.

## Funding

FARF -A138898 to P.S.R. U01NS120836 to M.R.W., S.J.P., J.M.G., A.J.G., K.C.Z., B.R.S., D.P., Y.M., S.S., I.T., C.T., A.N., and P.S.R.

## Competing Interests

M.R.W. has received unrelated research grant funding from Roche/Genentech, Kyverna Therapeutics and Novartis and is a co-founder and board member of Delve Bio, Inc. He has received consulting fees from Pfizer, Ouro Medicines, Vertex Pharmaceuticals and Indapta Therapeutics. The other authors declare no competing interests to the work of this manuscript.

## Ethics Approval

All research was approved at local institutional review boards and include appropriate clause for data sharing

## Data Availability

Given the rare nature of this disease and in order to protect privacy of the patients data sharing will be available only with permission.

## Author Contributions

MRW, GW, PSR, BS, and AN contributed to the conception and design of the study. GW, PSR, DP, AG, AA, NR, KZ, AS, YM, IT, SP, NG, and CT contributed to the acquisition and analysis of data. GW, DP, PSR, KZ, MRW, CL, CF, and RGM drafted a significant portion of the manuscript or figures (i.e., a substantial contribution beyond copy editing).

## AI Statement

Generative AI was used to edit the manuscript to improve readability and aid with drafting.

## Notes

### Competing Interest Statement

The authors have declared no competing interest.

### Author Declarations

UCSF IRB 13-12236

## References

1. Moreno O, Paredes A, Suarez-Obando F, Rojas A. An update on Fanconi anemia: Clinical, cytogenetic and molecular approaches (Review). Biomed Rep 2021;15(3):74.

2. Nalepa G, Clapp D. Fanconi anaemia and cancer: an intricate relationship. Nat Rev Cancer 2018;18(3):168–85.

3. Repczynska A, Julga K, Skalska-Sadowska J. Next-generation sequencing reveals novel variants and large deletion in FANCA gene in Polish family with Fanconi anemia. Orphanet J Rare Dis 2022;17(1):282.

4. Hoover A, Turcotte LM, Phelan R, et al. Longitudinal clinical manifestations of Fanconi anemia: A systematized review. Blood Reviews 2024;68:101225.

5. Stivaros SM, Alston R, Wright NB, et al. Central nervous system abnormalities in Fanconi anaemia: patterns and frequency on magnetic resonance imaging. Br J Radiol 2015;88(1056):20150088.

6. Aksu T, Gumruk F, Bayhan T, Coskun C, Oguz K, Unal S. Central nervous system lesions in Fanconi anemia: Experience from a research center for Fanconi anemia patients. Pediatr Blood Cancer 2020;67(12).

7. Bahar I, Weinberger D, Kramer M, Axer-Siegel R. Retinal vasculopathy in Fanconi anemia: a case report. Retina 2005;25(6):799–800.

8. Bartlett A, Wagner J, Jones B. Fanconi anemia neuroinflammatory syndrome: brain lesions and neurologic injury in Fanconi anemia. Blood Adv 2024;8(12):3027–37.

9. Chai S, Mathur R, Ong S. Retinal vasculopathy in Fanconi anemia. Ophthalmic Surg Lasers Imaging 2009;40(5):498–500.

10. Cousyn L, Demeret S, Philippi A. Autosomal recessive systemic microangiopathy associated with FANCL Fanconi anaemia. J Neurol Neurosurg Psychiatry 2023;95(1):98–100.

11. Denny M, Haug S, Cunningham EJ, Jumper J. Fanconi Anemia Presenting as Bilateral Diffuse Retinal Occlusive Vasculopathy. Retin Cases Brief Rep 2016;10(2):171–4.

12. Gayatri N, Hughes M, Lloyd I, Wynn R. Association of the congenital bone marrow failure syndromes with retinopathy, intracerebral calcification and progressive neurological impairment. Eur J Paediatr Neurol 2002;6(2):125–8.

13. Iki S, Shimizu H, Morimoto Y. Intracranial calcification and psychotic symptoms after irradiation in a patient with Fanconi anemia: A case report. PCN Rep 2022;1(2):e10.

14. Nathoo N, Gavrilova R, Trejo-Lopez J. Recurrent Tumefactive Central Nervous System Lesions Due to BRIP1 -Related Fanconi Anemia. Neurologist 2023;28(5):332–4.

15. Niederer RL, Ma SP, Wilsher ML, et al. Systemic Associations of Sarcoid Uveitis: Correlation With Uveitis Phenotype and Ethnicity. Am J Ophthalmol 2021;229:169–75.

16. Yahia S, Touffahi S, Zeghidi H, Zaouali S, Khairallah M. Ocular neovascularization in a patient with Fanconi anemia. Can J Ophthalmol 2006;41(6):778–9.

17. Hedderich DM, Lummel N, Deschauer M, et al. Magnetic Resonance Imaging Characteristics of Retinal Vasculopathy with Cerebral Leukoencephalopathy and Systemic Manifestations. Clin Neuroradiol 2020;30(2):229–36.

18. Kim H, de Jesus AA, Brooks SR, et al. Development of a Validated Interferon Score Using NanoString Technology. J Interferon Cytokine Res 2018;38(4):171–85.

19. Assarsson E, Lundberg M, Holmquist G, et al. Homogenous 96-plex PEA immunoassay exhibiting high sensitivity, specificity, and excellent scalability. PLoS One 2014;9(4):e95192.

20. Abdelhak A, Solomon I, Montes SC, et al. Retinal arteriolar parameters as a surrogate marker of intracranial vascular pathology. Alzheimer’s & Dementia: Diagnosis, Assessment & Disease Monitoring 2022;14(1):e12338.

21. Mayo ZS, Billena C, Suh JH, Lo SS, Chao ST. The dilemma of radiation necrosis from diagnosis to treatment in the management of brain metastases. Neuro Oncol 2024;26(12 Suppl 2):S56–65.

22. Stam AH, Kothari PH, Shaikh A, et al. Retinal vasculopathy with cerebral leukoencephalopathy and systemic manifestations. Brain 2016;139(11):2909–22.

23. Wilms AE, de Boer I, Terwindt GM. Retinal Vasculopathy with Cerebral Leukoencephalopathy and Systemic manifestations (RVCL-S): An update on basic science and clinical perspectives. Cereb Circ Cogn Behav 2022;3:100046.

24. Chauvin S, Ando S, Holley J. Inherited C-terminal TREX1 variants disrupt homology-directed repair to cause senescence and DNA damage phenotypes in Drosophila, mice, and humans. Nat Commun 2024;15(1):4696.

25. Khan M, Zhao Z, Arooj S, Liao G. Bevacizumab for radiation necrosis following radiotherapy of brain metastatic disease: a systematic review & meta-analysis. BMC Cancer 2021;21(1):167.

26. Dejonckheere CS, et al. Boswellia serrata for the management of radiation-induced cerebral edema and necrosis: a systematic meta-narrative review of clinical evidence. Adv Radiat Oncol 2025;10(4):101694.

1. Aksu T, Gumruk F, Bayhan T, Coskun C, Oguz KK, Unal S. Central nervous system lesions in Fanconi anemia: Experience from a research center for Fanconi anemia patients. Pediatr Blood Cancer. Dec 2020;67(12):e28722. doi:10.1002/pbc.28722

2. Bahar I, Weinberger D, Kramer M, Axer-Siegel R. Retinal vasculopathy in Fanconi anemia: a case report. Retina. Sep 2005;25(6):799–800. doi:10.1097/00006982-200509000-00023

3. Bartlett AL, Wagner JE, Jones B, et al. Fanconi anemia neuroinflammatory syndrome: brain lesions and neurologic injury in Fanconi anemia. Blood Adv. Jun 25 2024;8(12):3027–3037. doi:10.1182/bloodadvances.2024012577

4. Chai SM, Mathur R, Ong SG. Retinal vasculopathy in Fanconi anemia. Ophthalmic Surg Lasers Imaging. Sep-Oct 2009;40(5):498–500. doi:10.3928/15428877-20090901-11

5. Cousyn L, Demeret S, Philippi A, et al. Autosomal recessive systemic microangiopathy associated with FANCL Fanconi anaemia. J Neurol Neurosurg Psychiatry. Dec 14 2023;95(1):98–100. doi:10.1136/jnnp-2023-331260

6. Denny M, Haug SJ, Cunningham ET, Jr., Jumper JM. Fanconi Anemia Presenting as Bilateral Diffuse Retinal Occlusive Vasculopathy. Retin Cases Brief Rep. Spring 2016;10(2):171–4. doi:10.1097/ICB.0000000000000219

7. Gayatri NA, Hughes MI, Lloyd IC, Wynn RF. Association of the congenital bone marrow failure syndromes with retinopathy, intracerebral calcification and progressive neurological impairment. Eur J Paediatr Neurol. 2002;6(2):125–8. doi:10.1053/ejpn.2001.0559

8. Iki S, Shimizu H, Morimoto Y, et al. Intracranial calcification and psychotic symptoms after irradiation in a patient with Fanconi anemia: A case report. PCN Rep. Jun 2022;1(2):e10. doi:10.1002/pcn5.10

9. Nathoo N, Gavrilova RH, Trejo-Lopez JA, et al. Recurrent Tumefactive Central Nervous System Lesions Due to BRIP1 -Related Fanconi Anemia. Neurologist. Sep 1 2023;28(5):332–334. doi:10.1097/NRL.0000000000000511

10. Niedermayer I, Reiche W, Graf N, Mestres P, Feiden W. Cerebroretinal vasculopathy and leukoencephalopathy mimicking a brain tumor. Report of two early-onset cases with Fanconi’s anemia-like phenotypes suggesting an autosomal-recessive inheritance pattern. Clin Neuropathol. Nov-Dec 2000;19(6):285–95.

11. Yahia SB, Touffahi SA, Zeghidi H, Zaouali S, Khairallah M. Ocular neovascularization in a patient with Fanconi anemia. Can J Ophthalmol. Dec 2006;41(6):778–9. doi:10.3129/i06-078

12. Huang Y, Song C, He J, Li M. Research progress in endothelial cell injury and repair. Front Pharmacol. 2022;13:997272. doi:10.3389/fphar.2022.997272

13. Wang P, Konja D, Singh S, Zhang B, Wang Y. Endothelial Senescence: From Macro-to Micro-Vasculature and Its Implications on Cardiovascular Health. Int J Mol Sci. Feb 6 2024;25(4)doi:10.3390/ijms25041978

