## Supplemental Material for "Neuro-Ocular Vasculopathy Associated with Fanconi Anemia: A Clinical-Radiologic Phenotypic Case Series"

Supplemental Methods:

***Metagenomic Next Generation Sequencing***

These methods have been described previously but adjusted for the specifics of this experiment here.[1] Total nucleic acid was extracted from 2.5 mL of plasma of patients using the PAXgene Blood RNA Kit Handbook (Catalog No. 762174) and 90ul of CSF via the QIAGEN QIAcube. RNA-seq libraries were prepared using the New England Biolabs' NEBNext Ultra II RNA library preparation kit (Catalog No. E7770; NEB) as per the manufacturer's protocol. Library preparation was performed in bulk per protocol using the Echo Labcyte 225 and Agilent Bravo or Integra liquid handling robots.[2] Host ribosomal RNA depletion was performed using the Qiagen QIAseq FastSelect RNA removal kit (Catalog No. 333180; Qiagen) at 1:100 dilution. Samples were then amplified with incorporation of dual, unique indexing primers.[3] After pooling the dual-indexed libraries, the pool was size-selected using Ampure beads. The concentration was determined using a Kapa Universal quantitative PCR kit. The samples were then sequenced on an Illumina NovaSeq 6000 instrument using 150-base pair paired-end sequencing.

Sequences were uploaded to CZID, an open-source cloud-based metagenomics analysis pipeline.[4] The pipeline consists of iterative steps of sequence removal, beginning with the removal of human sequences through comparison to the most recent draft of the human genome. The remaining, nonhuman reads are searched in an indexed version of the National Center for Biotechnology Information nucleotide and nonredundant protein databases to identify the source(s) of the nonhuman sequences.

***Luminex cytokine analysis:***

Luminex - EMD Millipore Human 48 Plex Kits Protocol This assay was performed by the Human Immune Monitoring Center at Stanford University - Immunoassay Team. Kits were purchased from EMD Millipore Corporation, Burlington, MA, and run according to the manufacturer's recommendations with modifications described as follows: The H48 kits include one panel: Milliplex HCYTA-60K-PX48. The assay setup adhered to the recommended protocol with the following steps: 1. Sample Preparation: Samples were diluted 3-fold in a 96-well plate. (Plasma) Supernatant samples we ran undiluted. 2. Incubation: The samples were incubated overnight at 4°C with shaking. Cold and room temperature incubation steps were performed on an orbital shaker at 500-600 rpm. 3. Washing: Plates were washed twice with wash buffer using a BioTek ELx405 washer (BioTek Instruments, Winooski, VT). 4. Detection: After a one-hour incubation at room temperature with a biotinylated detection antibody, streptavidin-PE was added and incubated for 30 minutes with shaking. 5. Final Wash and Reading: Plates were washed as described above, and Wash Buffer was added to the wells for reading in the Luminex FlexMap3D Instrument, ensuring a lower bound of 50 beads per sample per cytokine. 6. Replication and Quality Control: Each sample was measured in duplicate replicates. Custom Assay Chex control beads (Radix BioSolutions, Georgetown, Texas) were added to all wells. Wells with a bead count <50 were flagged, and data with a bead count <20 were excluded. This method ensures precise and reliable quantification of cytokines using the Luminex FlexMap3D system.

***Type 1 interferon-score:***

The methods have been described previously and are summarized here.[5] Total RNA was extracted from blood samples collected in PAXgene Blood RNA tubes (Qiagen, Germantown, MD). Transcriptional analysis of 28 type I interferon-response genes (IRGs) was determined by NanoString (NanoString Technologies, Seattle, WA). A 28-gene type I IFN score was calculated as previously described, defined as the sum of the z-scores of 28 IRGs. Individual gene z-scores were calculated using the mean and standard deviation of gene counts from healthy controls.

***Rodent Brain Immunotaining:***

Thes methods have been described previously, and summarized again, here.[6] Adult, C57B6 male mice were perfused with 4% PFA and the brain was subsequently dissected and cryopreserved in 20% sucrose. Fixed brains were embedded in OCT, slowly frozen on dry ice and stored at -80 until needed. Fixed brains were sectioned on a cryostat at -20 in 12 micron sections onto superfrost plus microscope slides. Sections were dried for 1 hour at RT and then stained with antibodies according to the Immunocytochemistry (ICC) staining protocol described below. ICC and IHC experiments were visualized using a Nikon Ti Spinning Disc confocal microscope. Image capture settings, including exposure time, laser intensity, aperature, magnification were kept constant for all conditions in the experiment. Image TIFFs were analyzed in Image J and colocalization of RFP and GFP fluorescence was qualitatively determined.

ICC Staining protocol: Following washing with 1X PBS, cells were permeabilized and blocked for one hour at room temperature in Blocking buffer (1X PBS, 10% Goat Serum, 0.1% Triton). Following blocking, cells were incubated in one mL of primary antibody buffer (X PBS, 10% Goat Serum, 0.1% Triton) containing one of the following: Patient IgG (CSF 1:1000, Serum 1:10,000), commercial anti-KLH11 IgG (1 ug/mL) or commercial anti-FLAG IgG (1 ug/mL). Primary blocking buffer without antibody added was used as a secondary only control. Cells were incubated in primary antibody for 2 hours at room temperature or 4 degrees overnight. Cells were washed 3X in PBST (1X PBS with 0.1% Triton-X) and incubated with secondary antibodies for 1 hour at room temperature, protected by light. To detect human antibodies, antiHuman IgG Alexa-568 was used. To detect rabbit or mouse commercial antibodies, anti-Rabbit IgG or Anti-Mouse IgG Alexa 488 was used. Following secondary antibody incubation, cells 5 were washed 4X in PBST and mounted on coverglass for microscopy. DAPI was added during mounting using standard DAPI Fluormount-G (Southern Biotech).

***CSF protein Abundance (Olink)***

Clinically collected, neat CSF samples were selected from consenting patients with NOVA-FA (n=3) and from biopsy confirmed primary central nervous system small vessel vasculitis (n=3). The patients The three patients with biopsy confirmed CNS Vasculitis, were all female, ages at the time of CSF collection were 22,32, and 42, and race reported as White, Mixed (Black, White and Other, and Black-African American, respectively. Over 5000 proteins were measured using the Olink Explore HT, proximity extension assay platform, presented results in Normalized Protein eXpression (NPX) units.[7] Linear regression models of protein abundance values after removal of outliers based on Cooks distance (>4/n) and robust z-scores (>3.5), were used to estimate the log fold changes in NOVA-FA samples compared to CNS vasculitis. Given the small sample size only univariate regressions were fit. P-values were adjusted for multiple comparisons using the Benjamini–Hochberg method. Gene Set Enrichment Analysis identified Gene Ontology pathways enriched in NOVA-FA compared to primary CNS Vasculitis, using ClusterProfiler.[8]

Supplemental References:

1. Karalius MC, Ramachandran PS, Wapniarski A, et al. Infection in Childhood Arterial Ischemic Stroke: Metagenomic Next-Generation Sequencing Results of the VIPS II Study. Stroke [Internet] [cited 2025 May 8];0(0). Available from: https://www.ahajournals.org/doi/10.1161/STROKEAHA.124.050548

2. Mayday MY, Khan LM, Chow ED, Zinter MS, DeRisi JL. Miniaturization and optimization of 384-well compatible RNA sequencing library preparation. PLOS ONE 2019;14(1):e0206194.

3. Wilson MR, Fedewa G, Stenglein MD, et al. Multiplexed Metagenomic Deep Sequencing To Analyze the Composition of High-Priority Pathogen Reagents. mSystems 2016;1(4):10.1128/msystems.00058-16.

4. Kalantar KL, Carvalho T, de Bourcy CFA, et al. IDseq—An open source cloud-based pipeline and analysis service for metagenomic pathogen detection and monitoring. GigaScience 2020;9(10):giaa111.

5. Kim H, de Jesus AA, Brooks SR, et al. Development of a Validated Interferon Score Using NanoString Technology. J Interferon Cytokine Res 2018;38(4):171–85.

6. Mandel-Brehm C, Dubey D, Kryzer TJ, et al. Kelch-like Protein 11 Antibodies in Seminoma-Associated Paraneoplastic Encephalitis. New England Journal of Medicine 2019;381(1):47–54.

7. Assarsson E, Lundberg M, Holmquist G, et al. Homogenous 96-plex PEA immunoassay exhibiting high sensitivity, specificity, and excellent scalability. PLoS One 2014;9(4):e95192.

8. Guangchuang Yu, Li-Gen Wang, Yanyan Han and Qing-Yu He. clusterProfiler: an R package for comparing biological themes among gene clusters. OMICS: A Journal of Integrative Biology. 2012, 16(5):284-287

### Supplemental Table 1: Details of Demographic, Clinical, Radiological and CSF Findings for all six patients diagnosed with NO-VA

| FA GENE<br>mRS | RACE/ETHNICITY | AGE RANGE OF ONSET, NOVA (YEARS) | INITIAL CLINICAL NOVA PRESENTATION | PRECEDING NEUROLOGICAL HISTORY | NEUROLOGICAL COURSE | SPINAL CORD INVOLVEMENT (SPINAL LEVEL) | SEIZURES | INITIAL NOVA MRI BRAIN | OPTHALMOLOGIC COURSE AND LAST VISUAL ACUITY | BRAIN BIOPSY RESULTS | BMT (AGE YEARS) | TIME FROM FA DIAGNOSIS TO NOVA (YEARS) | TIME FROM BMT TO NOVA (YEARS) | CSF RESULTS | JCV SERUM SEROLOGY | STERIOD SPARING AGENTS | YEARS OF FOLLOW UP FROM FANS DIAGNOSIS |
|---|---|---|---|---|---|---|---|---|---|---|---|---|---|---|---|---|---|
| FANCD-2<br><br>5 | White, Non Hispanic | 20-25 | Hyperacute left hemiparesis | 2 years of subacute gait difficulties and neurogenic bladder | Two acute episodes of Neurological deficits (hemiparesis, and encephalopathy) with stable interval deficits. First immediately after a physical trauma. | YES<br>(C5-C6) | No | Mass-like Enhancing Left Frontal Lesion with patchy diffusion restriction and mass effect | Retinal ischemia noted on routine exam followed by diagnosis of cataracts requiring surgery. Further evaluations were never performed.<br>OD: 20/50<br>OS:20/200 | NA | YES<br>(6) | 20 | 19 | Isolated Elevated Protein (>100 mg/dL)<br><br>Negative IgG Index, JCV PCR | Not Tested | None | 3 |
| FANCD-2<br><br>6 | White, Non Hispanic | 10-15 | Subacute right hand dysmetria and mild ataxia | Complex Migraine (headache and aphasia) | Progressive ataxia, neurocognitive and neuropsychiatric decline, and seizures.<br><br>One acute episode in the setting of a mass-like lesion leading to transition to hospice care and death. | YES<br>(T5) | YES | Multiple scattered T2 and contrast enhancing lesions. Well-formed lesions in Left corpus collosum. | Slow, progressive functionally significant visual loss bilaterally (OS worse than OD).<br><br>OD: 20/200<br>OS: <20/200<br><br>Formal Fluorescence Angiogram performed with evidence of retinal vasculopathy. | NA | YES<br>(8) | 7 | 6 | Isolated Elevated Protein (>100 mg/dL)<br><br>Negative IgG Index, JCV PCR | | IVIG<br>MMF<br>Plasmapheresis<br>Rituximab<br>Sirolimus<br>Tocilizumab | 11 |
| FANC-J<br><br>4 | White, Non Hispanic | 20-25 | Transient Hyperacute Left Hemiplegia | 2-3 months of clumsiness (dropping plates) | Progressive ataxia, hemiparesis, with seizures and mild cognitive deficits.<br><br>Two acute episodes of neurological deficits with interval progression.<br>First immediately after a physical trauma. | YES<br>(T6) | YES | Mass-like Enhancing Left Frontal Lesion with patchy diffusion restriction and mass effect, and multiple scattered punctate, enhancing lesions. | Mild visual loss bilaterally.<br><br>OD:20/40<br>OS:20/40 | Necrosis with abundant macrophages and vascular sclerosis. There were no findings of vasculitis. | NO | 19 | NA | | Negative | Anakinra<br>Sirolimus | 8 |
| FANC-F<br><br>6 | White, Non Hispanic | 10-15 | Subacute onset left hemiplegia, and LMN CN7 and CN6 palsy | 4 years of neurocognitive deficits impairing schoolwork.<br><br>1 year prior slurred speech and cranial nerve palsy in the setting of cyclosporin toxicity. | Progressive cognitive decline, ataxia and seizures, initially thought to be mediation toxicity.<br><br>One acute episode in the setting of increasing mass effect causing death. | YES<br>(T7-T8; leptomeningeal at the conus medullaris) | YES | Multiple scattered T2 and contrast enhancing lesions. Extensive hemosiderin deposition throughout b/l cortices. Mass-like Enhancing Lesion with mass effects and herniation. | Slow, progressive functionally significant vision loss bilaterally (OD worse than OS).<br>OD: 20/160<br>OS: 20/160 | Reactive gliosis with necrosis and calcification, perivascular lymphocytic infiltrate. Numerous macrophages, rare CD4/CD8 cells in the perivascular and parenchymal cells. | YES<br>(4.5) | 13 | 9 | Isolated Elevated Protein (<100 mg/dL)<br><br>Negative IgG Index, JCV PCR. | Unknown | Anakinra<br>Cyclosporin<br>IVIG | 7 |
| None Found<br><br>1 | Asian | 30-25 | Acute onset vertigo | 3 years prior acute onset, transient (72 hours) hemisensory loss | Stable intermittent vertigo, one episode of subacute R facial droop in the setting of corresponding enhancing lesion in the R facial colliculus. | NO | NO | Multiple scattered T2 and contrast enhancing lesions, predominately subcortical. | Slow, progressive functionally significant vision loss bilaterally (OS worse than OD). Diagnosed with retinal ischemia in second decade of life.<br><br>OD: 20/50<br>OS: Finger Counting Centrally<br><br>Formal Fluorescence Angiogram performed with evidence of retinal vasculopathy. | NA | NO | 9 | NA | Normal<br><br>Negative IgG Index | Not Tested | NA | 5 |
| FANC-C<br>3 | White, Non Hispanic | 20-25 | Recurrent episodes of transient vertigo (duration: seconds) | Prolonged vertigo after traumatic head strike | Progressive mild right hand dysmetria | NO | NO | Multiple scattered T2 and contrast enhancing lesions, predominately cerebellum and brainstem. | Cataracts diagnosed soon after bone marrow transplant requiring surgery.<br>OD: 20/40<br>OS:20/40<br><br>Formal Fluorescence Angiogram performed with evidence of retinal vasculopathy. | NA | YES<br>(6) | 23 | 17 | Isolated Elevated Protein (<100 mg/dL)<br><br>Negative IgG Index<br><br>Elevated CSF ACE | Not Tested | NA | 8 |

Supplemental Table 2: Qualitative Summary of Radiological Findings among all six patients diagnosed with NO-VA

| Location of Mass-like lesion with vasogenic edema | Associated Vasogenic Edema with Mass Effect | Punctate T2 and Enhancing Lesions | | | | SWI/Calcifications | | | | T1 Hypointensities of corpus collosum |
|---|---|---|---|---|---|---|---|---|---|---|
| | | Cortical | Subcortical | Brainstem | Cerebellum | Cortical | Subcortical | Brainstem | Cerebellum | |
| 1) Right Frontal<br>2) Right Anterior Corpus Collosum | Yes | - | - | + | - | + | - | + | - | + |
| 1) Left Anterior Corpus Collosum<br>2) Left Posterior Corpus Collosum | No | ++ | +++ | + | +++ | ++ | ++ | + | + | + |
| 1) Right Frontal<br>2) Left Frontal<br>3) Left Centrum Semiovale | Mixed | + | ++ | ++ | +++ | + | + | + | - | + |
| 1) Left Occipital<br>2) Left Parietal | Yes | + | + | + | + | ++ | ++ | + | - | + |
| None | NA | ++ | + | + | + | + | - | - | - | + |
| None | NA | + | + | + | ++ | - | - | - | - | - |

Supplemental Table 3: Pathology Results from Brain Biopsy in Two Patients living with NO-VA

| Pathology Report Patient 1 | Pathology Report Patient 2 |
|---|---|
| DIAGNOSIS: Brain mass, excision: Brain parenchyma with necrosis, some inflammation, and vascular wall thickening. See Note.<br>NOTE: Sections show brain parenchyma with necrosis. GFAP stain shows reactive gliosis. CD45 highlights few leukocytes. Per report, CD163 stain showed abundant macrophages, GMS and AFB, toxoplasma and SV40 stains are reported negative for microorganisms.<br>______<br>Per report, CD163 shows abundant macrophages. Olig 2 highlights reactive glials. Stains (GMS, AFB, SV40<br>and toxoplasma) are negative for microorganisms. PAS highlights positive material in the vessel walls. Ki-67<br>proliferation rate is low. VVG stain highlights a lack of elastic lamina around the hyalinized vessels. Trichrome staining highlights the thick-walled vessels and shows luminal narrowing and occlusion in the multiple small vessels. Viable tissue is not available on this material for further testing.<br>Tumor Adequacy Assessment<br>Material available adequate for molecular evaluation: No<br>Requires macrodissection: N/A<br>Estimated viable tumor fraction (%): N/A | DIAGNOSIS: Brain, resection: Brain parenchyma with necroinflammatory process, see note<br>NOTE: Histologic sections show glial tissue with reactive gliosis, with mild perivascular lymphocytic infiltrate and necrosis.<br>______<br>Three H&E slides reviewed.<br>Per report, immunohistochemical staining for JC, BK, and EB viruses are negative. Rare CD8+ cells are noted with significant CD4+ cells in both the perivascular spaces as well as the parenchyma. CD68<br>highlights numerous perivascular macrophages as well as intraparenchymal macrophages. Staining for BK virus shows minor cytoplasmic blush. Ki-67 highlights scattered cells showing nuclear positivity and likely highlights the aforementioned inflammatory infiltrate. LFB/PAS does not show definitive evidence of demyelination. SV-40 stain was performed inhouse which was negative.<br>Adequacy Assessment for Ancillary Testing<br>Material available adequate for molecular evaluation: N/A<br>Requires macrodissection: N/A<br>Estimated Fraction of Viable Lesional Cells (%): N/A |

Supplemental Table 4: Summary of research based testing results for patients consenting to biospecimen collection (n=3)

<table>
<tr><th></th><th>CSF mNGS</th><th>CSF Cytokine Analysis</th><th>CSF Neopterin</th><th>CSF Type 1 Interferon Score</th><th>CSF Rodent Staining</th></tr>
<tr><td>“Patient 1”</td><td rowspan="3">No Bacterial, Viral, Fungal or Parasite Organisms Identified</td><td>Not Performed</td><td>Not Performed</td><td>Not Performed</td><td>Weak nuclear staining in cortex layers 2/3-5. Cytoplasmic staining of Purkinje cells processes in molecular layer of cerebellum. Weak neuropil staining throughout the brain.</td></tr>
<tr><td>“Patient 2”</td><td>Elevated CSF IL-6 (Value: 17 Reference Range: <5)</td><td>Negative</td><td>Negative</td><td>Nuclear predominant staining, with some cytoplasmic staining. Pyramidal layer in hippocampus and accessory olfactory bulb positive.</td></tr>
<tr><td>“Patient 3”</td><td>Normal</td><td>Negative</td><td>Not Performed</td><td>Strong nuclear staining throughout brain</td></tr>
</table>

Supplemental Figure 1: Representative images of gadolinium enhancing mass-like lesions with and without surrounding edema from four patients living with NOVA

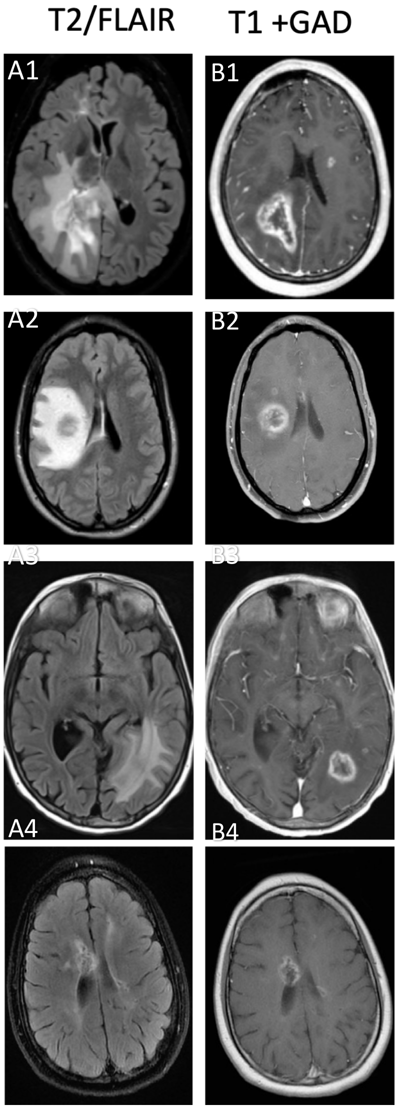


Patient presented with hyperacute, transient left hemiparesis and three months of left leg dragging, MRI Brain demonstrates rim enhancing right parietal lesion with surrounding edema (**A1**: Fluid attenuated inversion recovery (FLAIR) sequence; **B1**: gadolinium-enhanced $T_1$-weighted sequence). Different patient presenting with hyperacute left hemiparesis after 3 years of unexplained ataxia and bilateral leg stiffness,  MRI Brain demonstrates rim enhancing right frontal lesion with surrounding edema (**A2**: FLAIR sequence; **B2**: gadolinium-enhanced $T_1$-weighted sequence). Different patient with subacute onset left hemiplegia, and lower motor neuron cranial nerve six and seven palsy, MRI Brain demonstrates rim enhancing left parietal lesion with surrounding edema (**A3**: FLAIR sequence; **B3**: gadolinium-enhanced $T_1$-weighted sequence). Different patient presenting with subacute right hand weakness and difficulty with balance, MRI Brain demonstrates rim enhancing lesion in the right anterior corpus collosum without surrounding edema (**A4**: FLAIR sequence; **B4**: gadolinium-enhanced $T_1$-weighted sequence).
Abbreviations: FLAIR, Fluid-Attenuated Inversion Recovery , T1+ GAD $T_1$ Weighted Sequences with Gadolinium Enhancement

Supplemental Figure 2: Representative images of cervical and thoracic spinal cord lesions from three patients living with NOVA

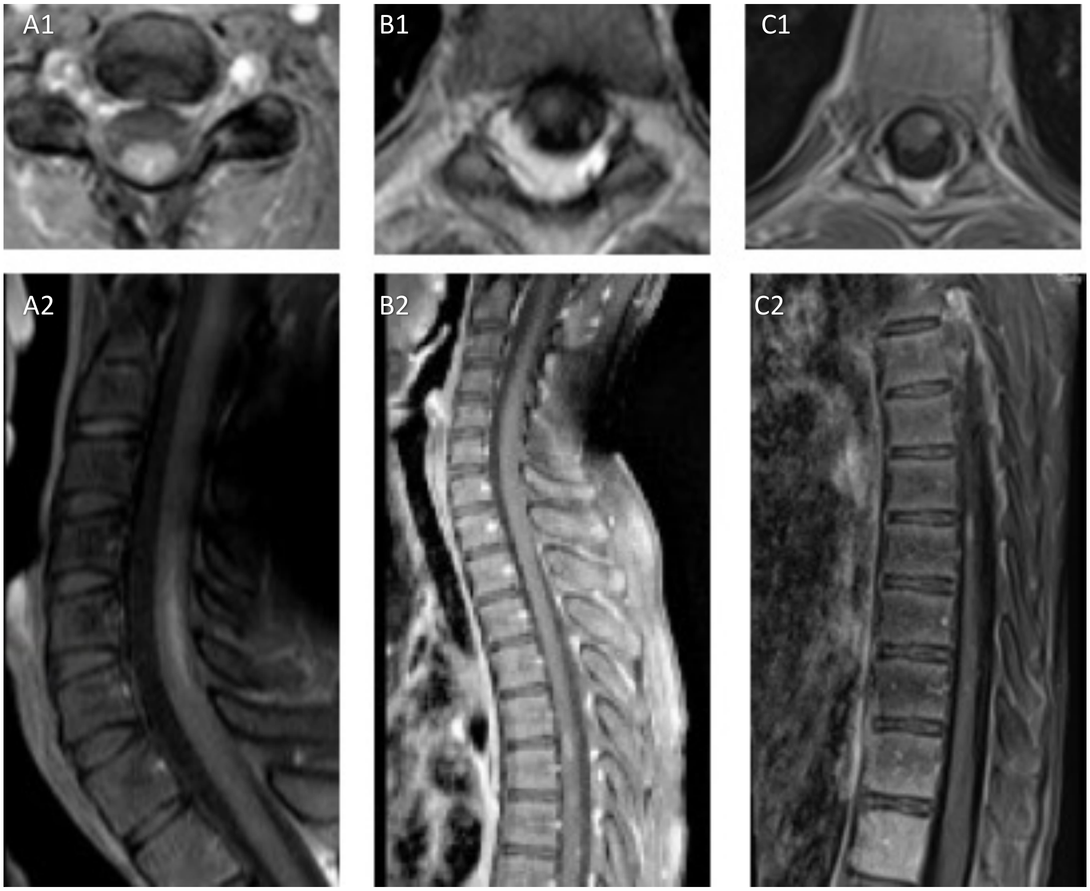


Patient presented with hyperacute left hemiparesis after 3 years of unexplained ataxia and bilateral leg stiffness, MRI Cervical Spine demonstrates a C5-6 intramedullary centrally enhancing lesion without surrounding edema (**A1**: axial gadolinium-enhanced $T_1$-weighted sequence; **A2**: sagittal gadolinium-enhanced $T_1$-weighted sequence). Different patient presenting with subacute right hand weakness and difficulty with balance, MRI Thoracic Spine demonstrates a T5 intramedullary, right, anterior gadolinium enhancing lesion without surrounding edema (**B1**: axial gadolinium-enhanced $T_1$-weighted sequence; **B2**: sagittal gadolinium-enhanced $T_1$-weighted sequence). A different patient with subacute onset left hemiplegia, and lower motor neuron cranial nerve six and seven palsy, MRI Thoracic Spine demonstrates T7 intramedullary anterior left gadolinium enhancing lesion without surrounding edema (**C1**: axial gadolinium-enhanced $T_1$-weighted sequence; **C2**: sagittal gadolinium-enhanced $T_1$-weighted sequence).

Supplemental Figure 3: Characterization of Pattern of Cerebral Atrophy

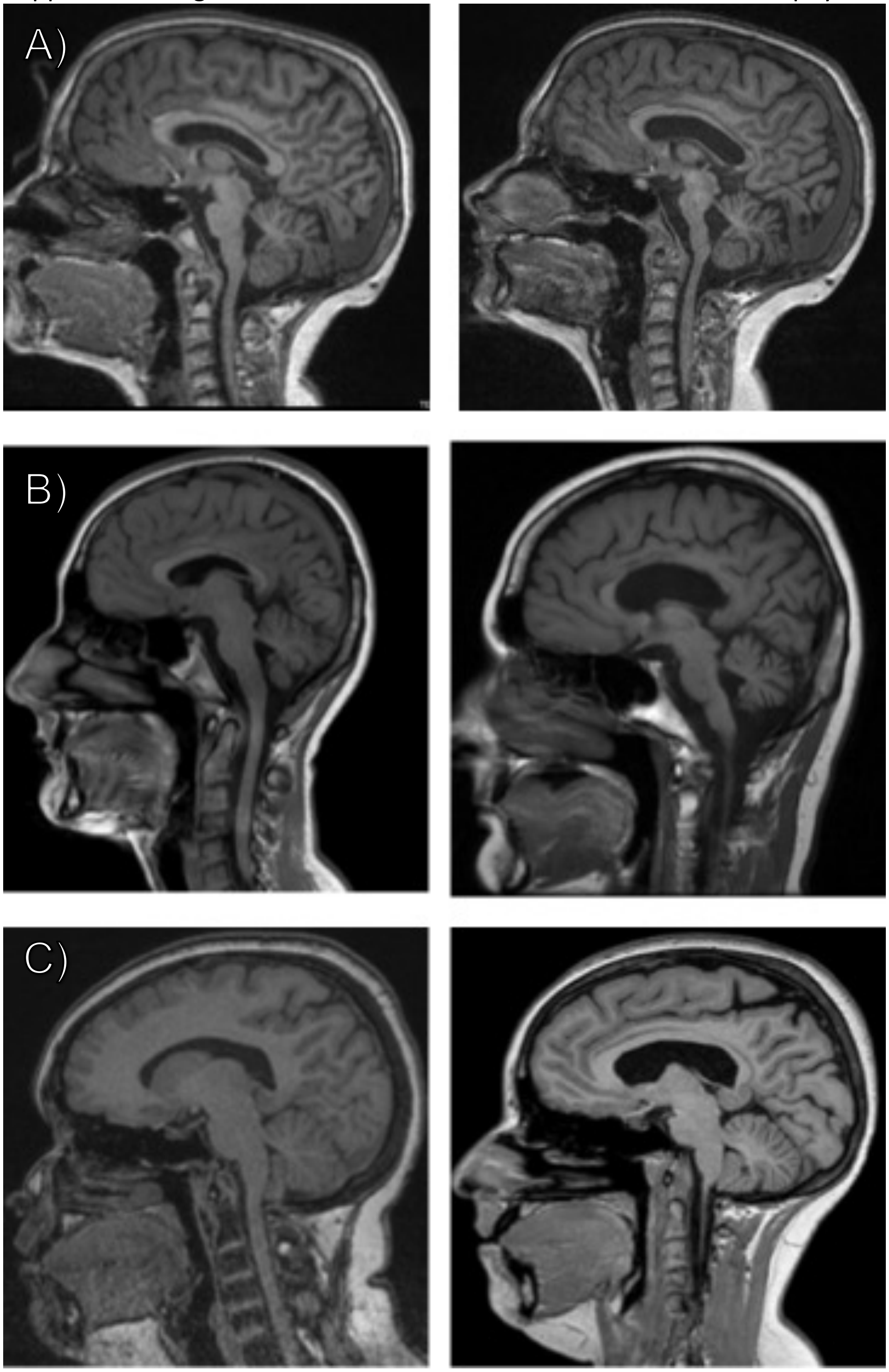


Sagittal T1-weighted images for three patients (A, B,C), respectively illustrating qualitative cerebral and cerebellar atrophy. Left hand columns reflect the first available sagittal Magnetic Resonance Image (MRI) and right column reflects the most recent image, all images are at least 3 years apart in calendar time.

Supplemental Figure 4: Representative images of mineral deposition among six patients living with NO-VA

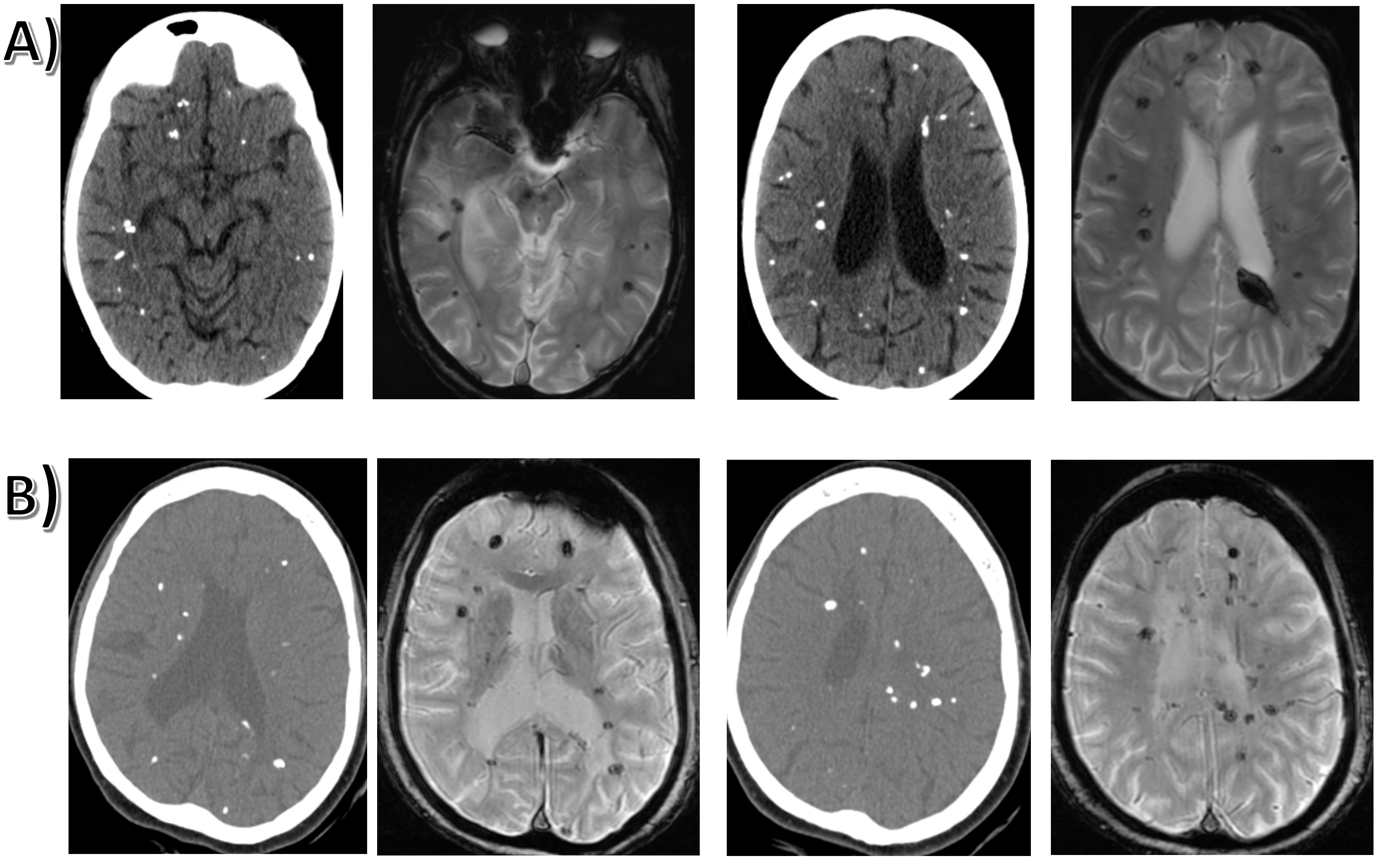


Representative images of corresponding CT images (columns 1 and 3) with MRI Susceptibility Weighted Images (columns 2 and 4) for two patients living with NO-VA (A) and B))

Supplementary Figure 5. Representative histologic findings from brain biopsy of a patient with NO-VA, Patient 1 as documented in Supplementary Table 3. Histopathologic examination of brain tissue revealed cerebral vascular narrowing and large sheets of parenchymal necrosis (A), abundant macrophages and reactive gliosis (B), and vessel hyalinization without evidence of transmural involvement by inflammatory cells (C-D). Immunohistochemical and special stains to further assess cerebrovascular pathology demonstrated thickened vascular walls on Trichrome (E) in vessels lacking internal elastic lamina on elastin stain (F). Thickened blood vessels also lacked mural PAS deposits (G). Abundant perivascular and intraparenchymal macrophages and microglia were highlighted in CD168 immunohistochemistry (H) but no granulomatous inflammation affecting vessels was noted.

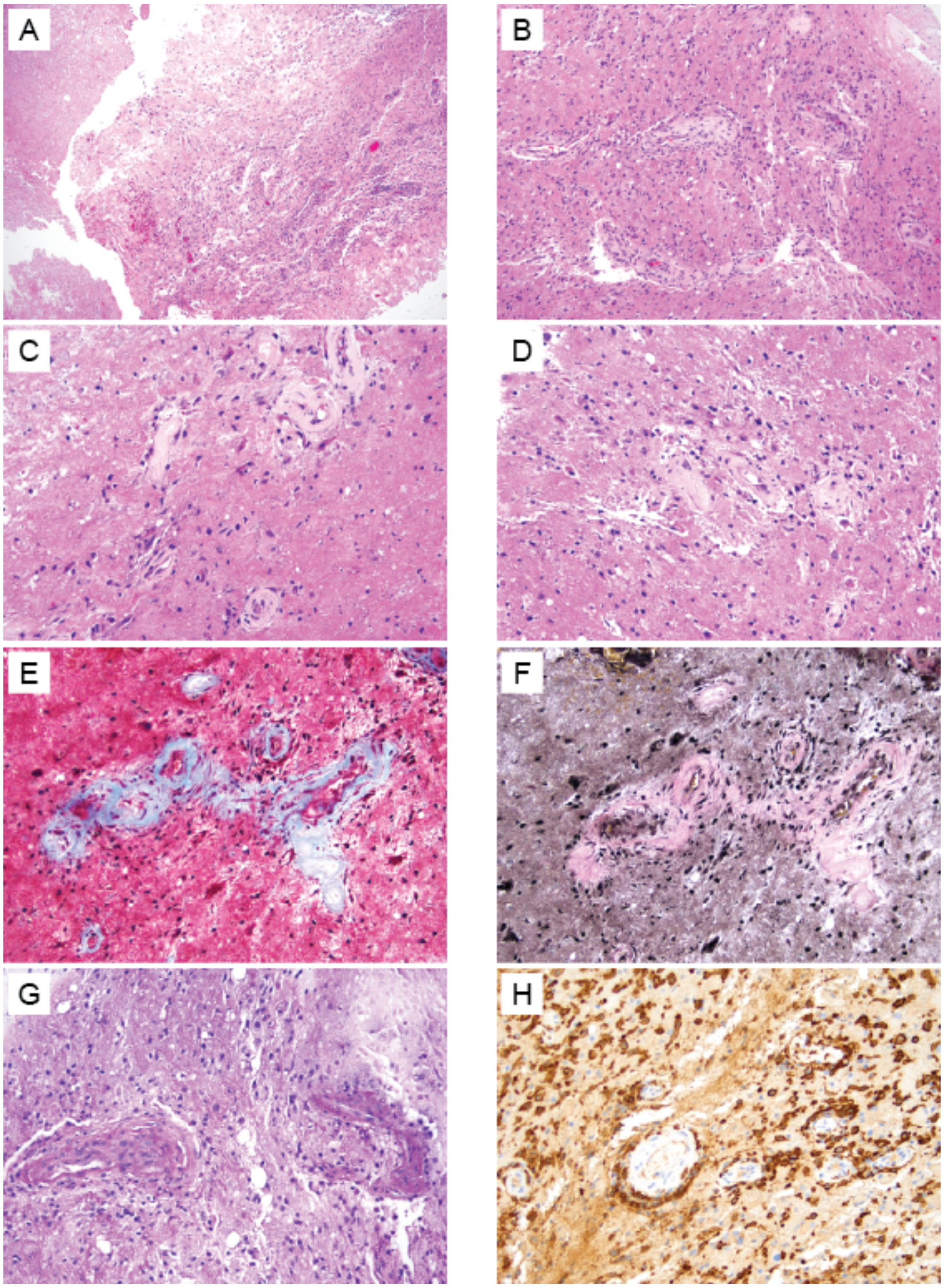

Supplemental Figure 6: Protein Abundance Differences between NOVA-FA (n=3) and biopsy-proven CNS Vasculitis (n=3). A) Volcano plot comparing protein abundance with significant proteins plotted in red, and top 20 (absolute log fold change) labeled. B) Log-fold change of all 118 statistically significant proteins.

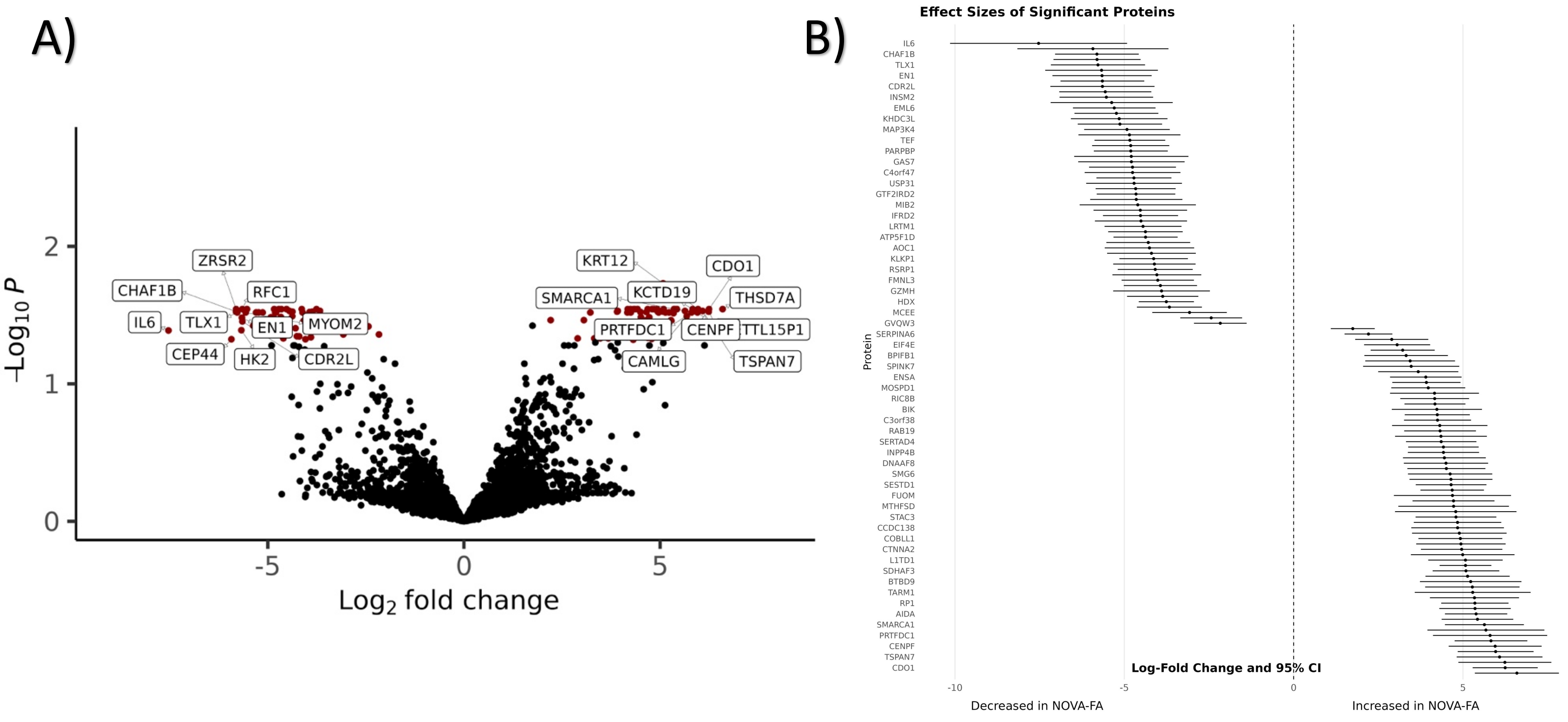

Supplemental Figure 7: Gene Set Enrichment Analysis identified Gene Ontology pathways enriched in NOVA-FA (n=3) compared to Primary CNS Vasculitis (n=3)

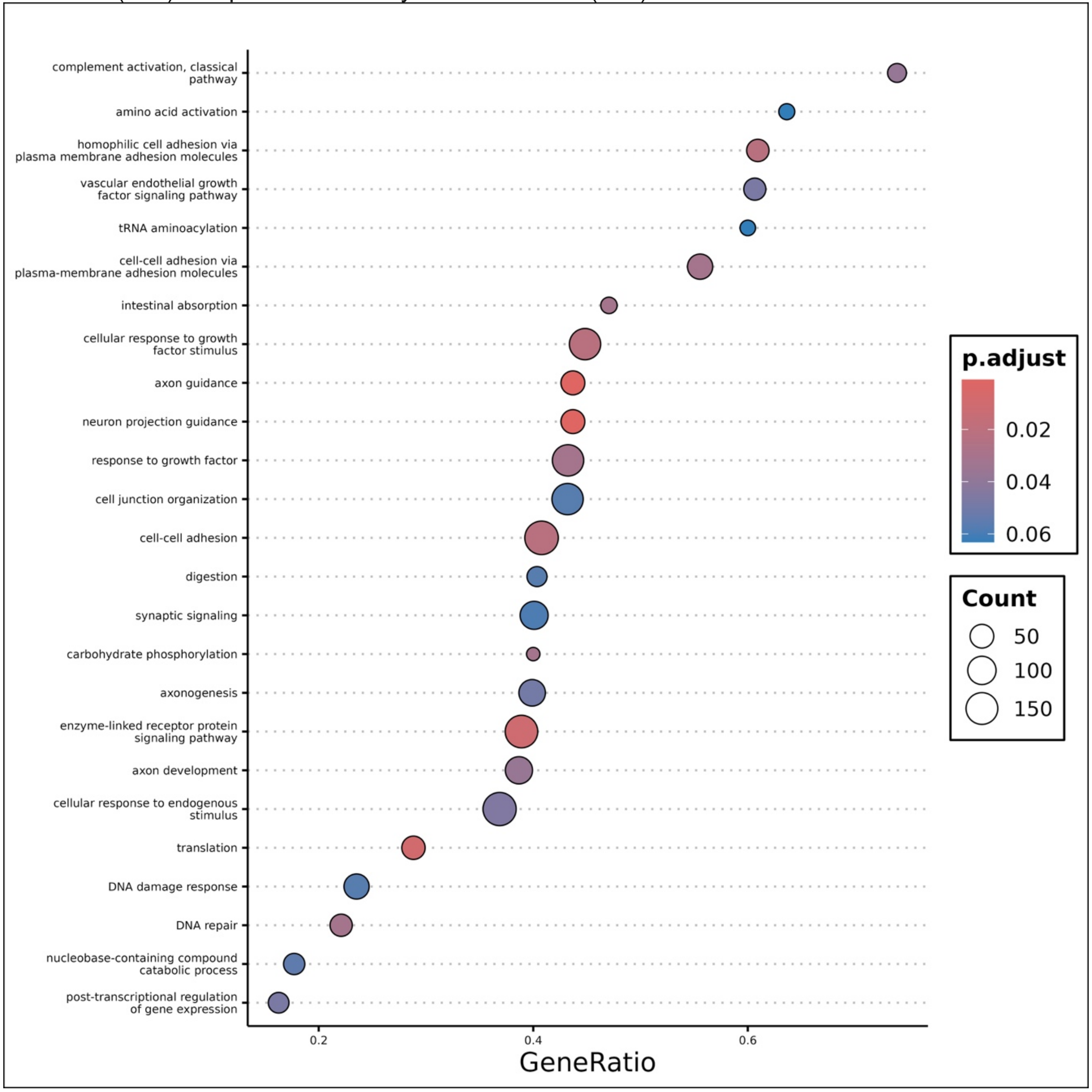

Supplemental Figure 8: Summary of clinical similarities and differences between NOVA, Cerebroretinal Vasculopathies

| | NOVA-FA | RVCL / HERNS / HVR |
|---|---|---|
| **Causal Genetic Variant** | ? | TREX-1 (c-terminus frameshift mutation) |
| **Age Onset** | 20s | Neurological Findings 50s |
| **MRI Brain Findings** | 1. Multiple punctate areas of T2 hyperintensity with enhancement in periventricular deep white matter, brainstem and cerebellum.<br>2. Post-contrast enhancing pseudotumor/space occupying lesions of variable size<br>3. Focal calcifications/SWI artifacts<br>4. Persistent Contrast Enhancement | 1. Multiple focal areas of T2 hyperintensity in periventricular and deep white matter<br>2. Post-contrast enhancing pseudotumor/space occupying lesions of variable size<br>3. Focal calcifications/SWI artifacts<br>4. Persistent Contrast Enhancement |
| **Neuroanatomical Lesions of Mass Lesions** | 1. Corpus Collosum<br>2. Frontal White Matter<br>3. Parietal White Matter | 1. Frontal White Matter<br>2. Periventricular deep grey and white matter<br>3. Cerebellum (in late stages) |
| **Spinal Cord Involvement** | YES | Not Reported |
| **Neurologic Presentation** | 1) Hyperacute deficits without resolution<br>2) Subacute and progressive<br>3) Seizures | Subacute the visual impairment, progressive blindness<br>Mild Neurological Symptoms until after age 50 which progress rapidly |
| **Associated Symptoms** | | Migraine, Raynaud's Phenomena, HTN |
| **CSF Findings** | Isolated elevated Protein<br>?Increased CSF IL-6 | No pleocytosis<br>Mildly elevated Protein |
| Retinal Fluorescein Angiogram | Decreased lumen diameter, telangiectasias, enlarged avascular fovea zone | capillary dropout, especially in the macula, leading to loss of central vision, telangiectasias and juxta-foveolar capillary obliteration |
| Brain Biopsy | Necrosis with abundant macrophages and vascular sclerosis, without evidence of vasculitis. | Confluent foci of coagulation necrosis surrounded by reactive gliosis, Focal calcifications<br>Fibrinoid vascular necrosis, adventitial fibrosis, luminal narrowing and mural hyalinization |
| **Response to Glucocorticoids** | None | Temporary Symptomatic Benefit |
| **RCT Treatment** | None | Aclarubicin |
| **Trialed Treatments (Off Label Clinical)** | Anakinra<br>Tocilizumab | Azathioprine, Bevacizumab (intravitreal), Chloroquine, Cyclophosphamide, Methotrexate |
| **Ongoing Treatment Trials** | None | Crizanlizumab |

Abbreviations: NOVA-FA, Neuro-Ocular Vasculopathy Associated with Fanconi Anemia, RVCL, Retinal Vasculopathy with Cerebral Leukodystrophy, HERNS, Hereditary Endotheliopathy, Retinopathy, Nephropathy, and Stroke HVR, Hereditary Vascular Retinopathy
